# Right For Me: a pragmatic multi-arm cluster randomised controlled trial of two interventions for increasing shared decision-making about contraceptive methods

**DOI:** 10.1101/2021.06.25.21257891

**Authors:** Rachel Thompson, Gabrielle Stevens, Ruth Manski, Kyla Z Donnelly, Daniela Agusti, Zhongze Li, Michelle Banach, Maureen B Boardman, Pearl Brady, Christina Colón Bradt, Tina Foster, Deborah J Johnson, Sarah Munro, Judy Norsigian, Melissa Nothnagle, Ardis L Olson, Heather L Shepherd, Lisa F Stern, Tor D Tosteson, Lyndal Trevena, Krishna K Upadhya, Glyn Elwyn

## Abstract

**Objectives:** There is a paucity of evidence on how to facilitate shared decision-making under real-world conditions and, in particular, whether interventions should target patients, health care providers, or both groups. Our objectives were to assess the comparative effectiveness, feasibility, and acceptability of patient- and provider-targeted interventions for improving shared decision-making about contraceptive methods in a pragmatic trial that prioritised applicability to real-world care.

**Design:** The study design was a 2×2 factorial cluster randomized controlled trial with four arms: (1) video + prompt card (‘video’), (2) decision aids + training (‘decision aids’), (3) dual interventions (‘dual’), and (4) usual care. Clusters were 16 primary and/or reproductive health care clinics that deliver contraceptive care in the Northeast United States.

**Participants:** Participants were people who had completed a health care visit at a participating clinic, were assigned female sex at birth, were aged 15-49 years, were able to read and write English or Spanish, and had not previously participated in the study. Participants were enrolled for 13 weeks before interventions were implemented in clinics (pre-implementation cohort) and for 26 weeks after interventions were implemented in clinics (post-implementation cohort). 5,018 participants provided data on at least one study outcome.

**Interventions:** Interventions were a video and prompt card that encourage patients to ask three specific questions in the health care visit and a suite of decision aids on contraceptive methods and training for providers in how to use them to facilitate shared decision-making with patients in the health care visit.

**Main outcome measures:** The primary outcome was shared decision-making about contraceptive methods. Secondary outcomes spanned psychological, behavioural, and health outcomes. All outcomes were patient-reported via surveys administered immediately, four weeks, and six months after the health care visit.

**Results:** We did not observe any between-arm difference in the differences in shared decision-making between the pre- and post-implementation cohorts for the sample as a whole (video vs. usual care: adjusted odds ratio (AOR)=1.23 (95% confidence interval (CI): 0.82 to 1.85), p=0.80; decision aids vs. usual care: AOR=1.47 (95% CI: 0.98 to 2.18), p=0.32; dual vs. video: AOR=0.95 (95% CI: 0.64 to 1.41), p=1.00; dual vs. decision aids: AOR=0.80 (95% CI: 0.54 to 1.17), p=0.72) or for participants with adequate health literacy. Among participants with limited health literacy, the difference in shared decision-making between the pre- and post-implementation cohorts was different in the video arm from the usual care arm (AOR=2.40 (95% CI: 1.01 to 5.71), p=.047) and was also different in the decision aids arm from the usual care arm (AOR=2.65 (95% CI: 1.16 to 6.07), p=.021), however these differences were not robust to adjustment for multiple comparisons. There were no intervention effects on the secondary outcomes among all participants nor among prespecified subgroups. With respect to intervention feasibility, rates of participant-reported exposure to the relevant intervention components were 9.4% for the video arm, 31.5% for the decision aids arm, and 5.0% for the dual arm. All interventions were acceptable to most patients.

**Conclusions:** The interventions studied are unlikely to have a meaningful population-wide impact on shared decision-making or other outcomes in real-world contraceptive care without additional strategies to promote and support implementation. Selective use of the interventions among patients with limited health literacy may be more promising and, if effective, could reduce disparities in shared decision-making.

**Trial registration:** ClinicalTrials.gov NCT02759939.

## BACKGROUND

Shared decision-making is the process of health care providers and patients making health decisions together. Although a number of conceptual definitions have been offered (1), the most common considers shared decision-making a process in which providers and patients exchange information, deliberate about available options together, and come to agreement on the option to implement (2). There is accumulating evidence suggestive of positive impacts of shared decision-making on patient psychological, behavioural and health outcomes in a range of clinical contexts (3). Shared decision-making may be a particularly worthwhile approach in the contraceptive care context. Due to its emphasis on knowing and respecting the preferences and values of each individual patient, shared decision-making may be a strategy for upholding patient autonomy and promoting health and wellbeing. For example, it may enable providers to facilitate the selection and use of effective contraceptive methods when this is the patient’s goal. It may also enable providers to identify when this is not the patient’s goal and tailor their counselling accordingly. Potential gains from shared decision-making in contraceptive care may be pronounced for people of low socioeconomic status and people from racial and ethnic minority groups in light of the historical context of coercive reproductive policies targeting them, enduring harms, and persisting disparities in contraceptive care and outcomes (4,5).

Despite observed and theoretical advantages of shared decision-making, there is limited evidence on facilitating its adoption in practice. First, it is uncertain whether implementation efforts should focus on patients, health care providers, or both groups simultaneously. A systematic review of interventions for improving adoption of shared decision-making that was published before this study began found that, although interventions targeting patients or providers alone had some—if modest—success, effects were generally greater when interventions targeted both groups (6). However, based on the small number and poor quality of available studies, reviewers were tentative in their conclusions and advocated further research addressing this question. Second, it is uncertain whether interventions that have previously shown promise for facilitating adoption of shared decision-making would prove useful under real-world conditions. In many studies in the systematic review referenced above, intervention strategies were reliant on resources not widely accessible in health care settings. In some, the intervention required a substantial investment of either time (e.g., a 20-hour communication skills course for hospital-employed doctors (7)) or financial resources (e.g., a strategy that included reimbursement of further health care in the event of an unfavourable decisional outcome (8)). In others, additional personnel were provided to undertake aspects of intervention implementation, such as providing an individualised briefing to health care providers immediately before seeing each patient (9).

We responded to this uncertainty by assessing the comparative effectiveness of patient- and provider-targeted interventions for improving shared decision-making about contraceptive methods in a pragmatic trial that prioritised applicability to real-world care. Our first objective was to assess the effect of a video and prompt card (patient-targeted intervention) and decision aids and training (provider-targeted intervention), delivered alone and together, on shared decision-making about contraceptive methods. Our second objective was to assess the effect of these interventions on various secondary psychological, behavioural, and health outcomes. Our third objective was to assess the feasibility of the patient-facing intervention components (operationalised as rates of participant exposure to them) and their acceptability to patients. Our specific research questions and hypotheses are provided in Appendix 1 and in our published study protocol (10).

## METHODS

The reporting of this study was guided by the CONSORT statement (11); CONSORT extensions for pragmatic trials (12), cluster randomised trials (13), multi-arm trials (14), trials with patient-reported outcomes (15), and trials of nonpharmacologic treatments (16); and the TIDieR checklist (17).

### Study design and overview

We conducted a 2×2 factorial cluster randomised controlled trial with four arms: (1) video + prompt card (‘video’), (2) decision aids + training (‘decision aids’), (3) video + prompt card and decision aids + training (‘dual’), and (4) usual care. The clusters were health care clinics and participants were patients with a health care visit at a participating clinic. We enhanced the conventional cluster randomised controlled trial design by enrolling patients receiving care during a 13-week period before interventions were introduced (the ‘pre-implementation cohort’) in addition to patients receiving care during a 26-week period after interventions were introduced (the ‘post-implementation cohort’) (18) (see Figure 1). Data were collected via patient surveys administered immediately (T1), four weeks (T2), and six months (T3) after the index health care visit. Ultimately, analyses compared the difference between the pre-implementation and post-implementation cohort between trial arms.

**Figure 1.**
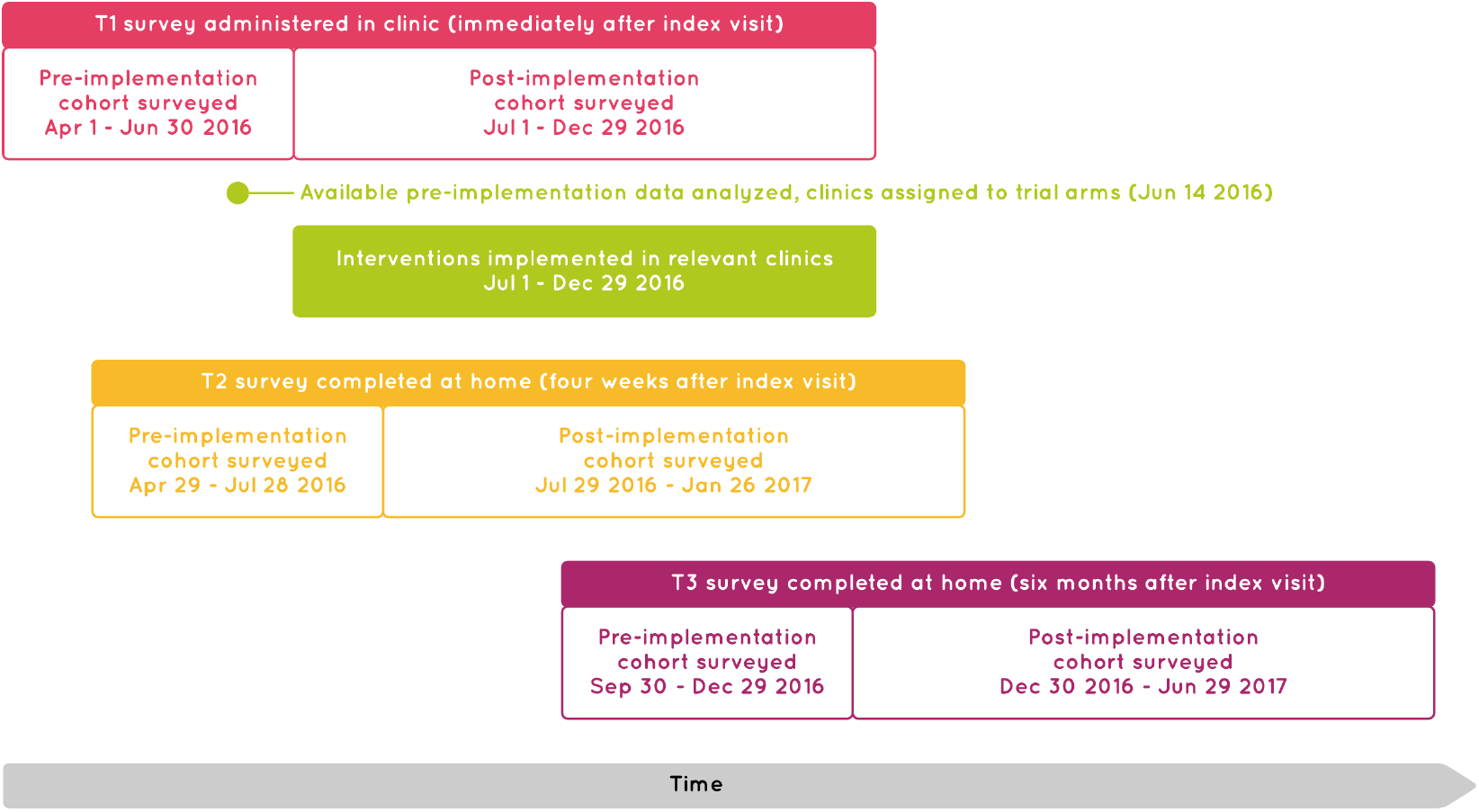
Study schematic

### Setting and clusters

The clusters were 16 clinics in the Northeast United States, meeting the recommended minimum of four clusters per trial arm to minimise potential confounding from cluster effects (13). Clinics had to deliver contraceptive care, defined as providing contraceptive methods on site, prescribing or referring people for contraceptive methods, and/or counselling people about contraceptive methods. To avoid contamination, clinics could not employ a study investigator and could not employ a person who delivered contraceptive care in another participating clinic. Additionally, clinics had to have sufficient patient flow for a representative to be confident that 10 eligible patients, on average, would enrol in the study each week. Clinics could be primary care and/or reproductive health care clinics and were not required to have any particular infrastructure or features. Within each clinic, contraceptive care could be provided by any person with relevant training (e.g., a physician, nurse, physician assistant, nurse practitioner, midwife, non-medically licensed counsellor). Clinics could operate in a hospital or community setting, be located in rural or urban areas, be publicly or privately funded, be for-profit or not-for-profit, and deliver services in more than one geographic location. Clinics were required to nominate at least one representative (of any position, discipline, or training) to liaise with the research team and coordinate study-related activities. Clinics received monetary compensation for the time dedicated to data collection and other study-related administration but not for the time and effort dedicated to implementing relevant interventions.

### Allocation

Each clinic was assigned to one of the four trial arms using permuted-block randomisation with an equal allocation ratio to achieve balance in the number of clinics per trial arm. We adopted stratified permuted-block randomization because, when we analysed available data from the pre-implementation cohort (on June 14, 2016; see Figure 1), we found non-equivalence among clinics in the rate of shared decision-making about contraceptive methods (clinic range: 35.8% to 73.2%, χ^2^=52.61, p<.001). We ranked clinics according to the rate of shared decision-making and constructed four strata based on this ranking, with one clinic assigned to each trial arm within each stratum. The study statistician (TT) and statistical analyst (ZL) generated the allocation sequence and undertook the assignment. Due to the study design, it was not feasible to blind the statistical analyst, other members of the research team, or clinic staff to the trial arm to which clinics were assigned. Participants were informed of the broad objectives of the study but were not privy to the specific hypotheses being tested.

### Participants

Principles of inclusivity guided our study eligibility criteria. Eligible to enrol in the trial were people who had completed a health care visit at a participating clinic, were assigned female sex at birth, were aged 15–49 years, were able to read and write English or Spanish, and had not previously participated. Given these broad eligibility criteria, further criteria were imposed for the assessment of some study outcomes to ensure their relevance and minimise participant burden (see Table A3 in Appendix 9). In addition, participants were eligible to give permission to be contacted for T2 and T3 data collection only if they reported at T1 that they had experienced a conversation about contraception during the health care visit and intended to use one or more contraceptive methods in the next four weeks. Participants were not required to have completed the T2 survey to be eligible for the T3 survey.

### Recruitment and consent

Research team members provided clinic staff with face-to-face training in recruitment and data collection processes at the beginning of the study. In some clinics, a face-to-face refresher was also provided around the time clinics were allocated to trial arms. Although tailoring of some recruitment and data collection processes to the clinic workflow was encouraged, most were uniform across clinics. People were made aware of the opportunity to participate in the study on the day of their health care visit via study posters and information sheets displayed in participating clinics (see Appendix 2) and/or through communication with clinic staff. Clinic staff provided potentially eligible patients with access to a tablet computer (proactively and/or on request) that displayed an electronic version of the study information sheet. This information sheet explained the study and enabled patients to self-assess their eligibility to enrol. Patients who selected ‘*Yes*’ when asked, ‘*Now that you have read this information, do you agree to participate in this study?*’ were considered to have provided informed consent to participate in data collection and immediately proceeded to the T1 survey. At the end of the T1 survey, participants eligible for T2 and T3 surveys were able to give or decline permission to be contacted about these surveys.

### Interventions and comparator

#### Video + Prompt Card (‘Video’)

The Right For Me video (https://vimeo.com/244655240, https://vimeo.com/244659975) was a brief video intended to be viewed by patients in the clinic before a health care visit. The video was designed to enhance patients’ motivation, skills, and self-efficacy to ask three questions in the health care visit: (a) what are my options? (b) what are the possible pros and cons of those options? and (c) how likely are each of those pros and cons to happen to me? Earlier iterations of these questions increased shared decision-making when used by unannounced standardised patients in a primary care setting in Australia (19) and were the focus of the AskShareKnow program, a multipronged question prompt intervention that was both feasible to implement in a reproductive and sexual health care setting in Australia and acceptable to patients (20). The Right For Me video was an adaptation of a 4-minute video that comprised one component of the AskShareKnow program.

The Right For Me video featured a patient sharing a personal account of her experiences of health care (with contraceptive care examples), communicating the benefits of asking questions, normalising discomfort with asking questions, and encouraging people to ask the three target questions. The format and content of the adaptation responded to patient and stakeholder perspectives solicited via discussions among the research team, listening sessions with patients, and consultation with the patient featured in the video, who later joined our research team. To allow for flexibility in implementation, we sought to maximise the salience of the video’s message for patients receiving contraceptive care without sacrificing its suitability for other populations. To enhance accessibility, we developed versions of the video in English (2:34 minutes) and Spanish (3:29 minutes), each with and without on-screen captions. We supplied clinics with two tablet computers programmed to view the video, two sets of headphones, and supplies for cleaning the tablet computers.

The Right For Me prompt card (see Appendix 3) was a wallet-sized card intended for patients who viewed the video. The card was designed to remind patients of the questions presented in the video and was adapted from a refrigerator magnet that was a second component of the AskShareKnow program (20). We supplied clinics with prompt cards in English and Spanish and display stands.

#### Decision Aids + Training (‘Decision aids’)

The Right For Me decision aids (see Appendix 4) were seven one-page decision aids on contraceptive methods intended to be used by providers with patients during the health care visit. There were decision aids on long-acting reversible contraceptive methods, short-acting reversible methods, barrier methods, natural methods, permanent methods, and emergency methods, as well a decision aid that provided an overview of these six categories of methods. The decision aids were designed to help providers facilitate shared decision-making about contraceptive methods in the health care visit. The format of the decision aids was adapted from Option Grid™ decision aids, which had been found to be acceptable to physicians (21) and increase shared decision-making in osteoarthritis care (22). We co-designed the decision aids with end users, including by determining patient and contraceptive care provider information priorities (23), holding listening sessions with patients to inform decision aid branding, language, and data presentation, and inviting feedback on drafts by both patient and stakeholder partners on the research team and stakeholders in our Advisory Group (see *Patient and public involvement*). We supplied clinics with paper tear-pads of the decision aids in English and Spanish and display stands.

The Right For Me training video (https://vimeo.com/244663383) and written guidance (see Appendix 5) were intended to be viewed by providers before beginning to use the decision aids and as often as needed thereafter. The training video and written guidance were informed by the Theoretical Domains Framework (24,25) and were designed to enhance providers’ motivation, skills and self-efficacy to use the decision aids to facilitate shared decision-making in the health care visit. The video (4:20 minutes) featured an obstetrician-gynaecologist, nurse practitioner, and patient representative explaining that decision aids can support the delivery of quality health care and providing guidance on using them. The written guidance reinforced and elaborated on training video content. We hosted the training video and written guidance in a password-protected area of the study website and asked clinic representatives to encourage and enable relevant health care providers in the clinic to review them.

#### Implementation of interventions

The strategies we developed to support implementation of the interventions in clinics purposefully omitted elements we considered infeasible or costly to scale up (e.g., face-to-face training, regular feedback on rates of intervention implementation). For both interventions, we developed a slide deck for clinic staff that provided guidance on intervention objectives and implementation but also encouraged tailoring to clinic features and processes where possible (see Appendices 6-7). The slide decks were hosted in password-protected areas of the study website. For clinics assigned to implement the video, previews of the video and prompt card accompanied the slide deck. Clinic representatives were asked to encourage and enable relevant staff members to review the slide deck(s). The interventions were to be implemented for a period of 26 weeks. To approximate real-world conditions, clinics were not asked to remove any usual resources and were permitted to continue or begin implementing concomitant interventions or care during the implementation period. Fidelity of intervention implementation was inferred from a combination of patient-reported data (see *Outcomes and measures*) and qualitative interviews with clinical and administrative staff upon completion of the trial (26).

#### Usual care

Clinics assigned to deliver usual care were not provided with either of the study interventions but were not prevented from implementing concomitant interventions or care.

### Time frame and data collection

We collected data via English and Spanish patient surveys administered at three time points: immediately after the health care visit (T1), four weeks after the health care visit (T2), and six months after the health care visit (T3) (surveys available on request). The T1 survey was administered electronically in clinics using tablet computers. This survey assessed outcomes including whether participants experienced a conversation about contraception in the health care visit and what, if any, contraceptive method(s) they intended to use in the next four weeks. The T2 and T3 surveys were completed by participants at home. Participants aged 20 years and older could elect to complete these surveys online (with correspondence via a supplied email address) or on paper (with correspondence via a supplied mailing address). Participants aged under 20 years could only elect to complete these surveys online (with correspondence via a supplied email address) to safeguard their privacy. A unique password linked participant responses across surveys.

As noted above, we enrolled participants during a 13-week period before interventions were introduced (the ‘pre-implementation cohort’) and during a 26-week period after interventions were introduced (the ‘post-implementation cohort’) (18). We had two reasons for enrolling the pre-implementation cohort: (1) to give clinic staff time to become comfortable with enrolment and data collection processes before potentially being asked to add intervention implementation to their workflow, and (2) to enable us to assess equivalence among clinics in the initial rate of shared decision-making about contraceptive methods (our primary outcome), adopt stratified assignment to trial arms if warranted, and account for pre-existing differences in our analyses. Ultimately, we made even greater use of data from the pre-implementation cohort (see *Analysis*).

### Outcomes and measures

Study outcomes and measures are listed in Table 1 and are further elaborated in Table A3 in Appendix 9. Although we prioritised patient-centered outcomes, we also included more conventional public health-oriented outcomes (e.g., intention to use a highly effective contraceptive method (27)) to maximise the value of our findings for diverse knowledge users. We selected measures based on their validity and reliability, brevity, readability, patient-centred tone and language, prior use, and availability in English and Spanish. When we identified a suitable measure in English that was not available in Spanish, we had it translated. When we could not identify a suitable measure, we developed or adapted one in English and had it translated into Spanish.

**Table 1.**
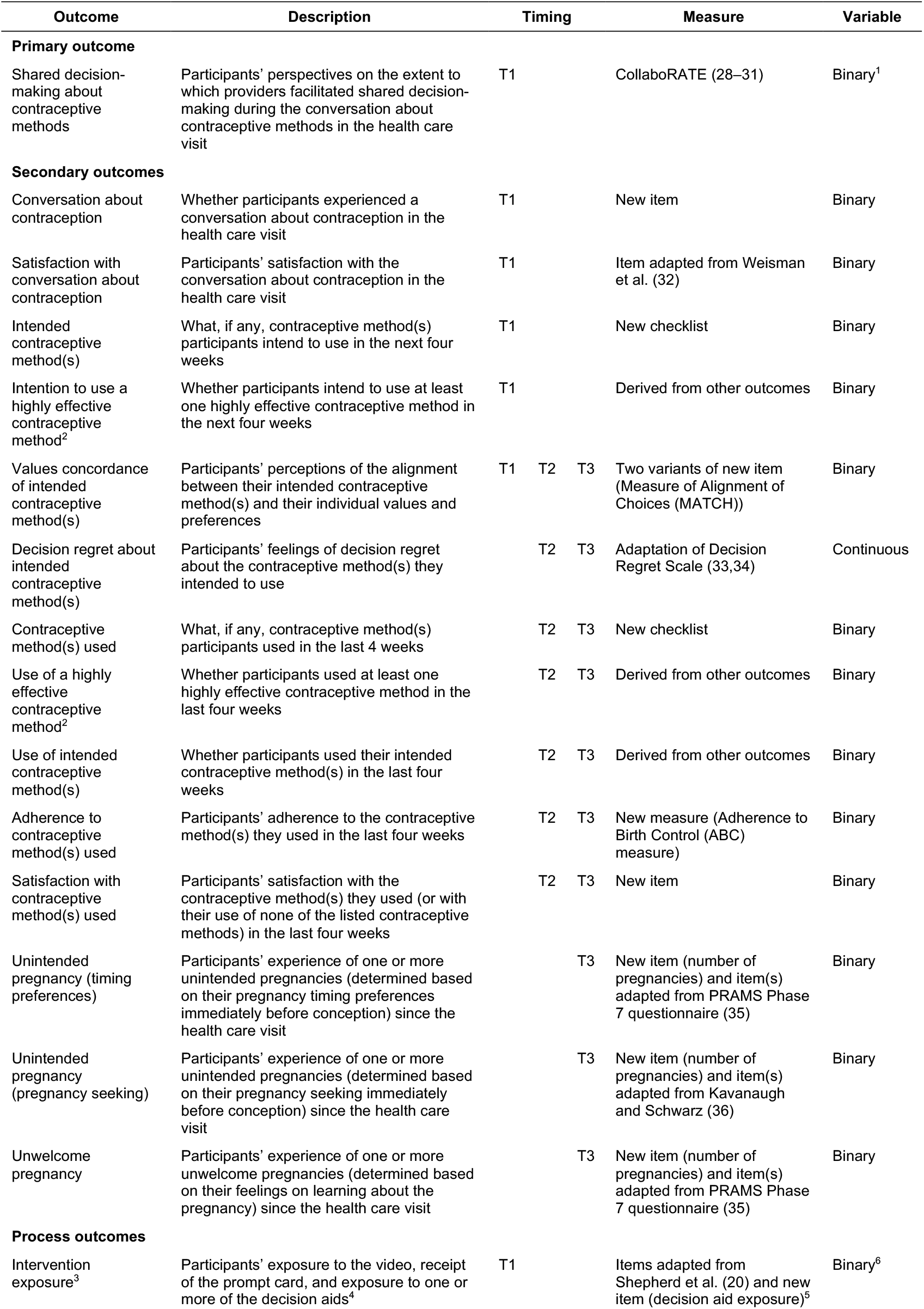
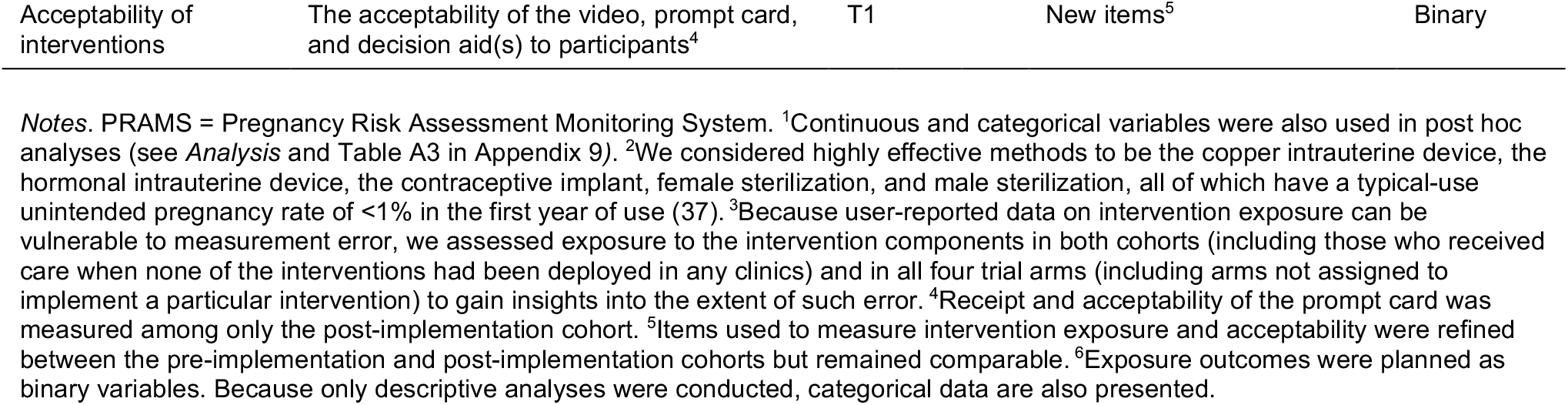
Outcomes and measures

The primary outcome – shared decision-making about contraceptive methods – was measured using CollaboRATE, which assesses patient perceptions of the extent to which a health care provider(s) shared information, elicited patient preferences, and ensured patient preferences were integrated as decisions were made (28–31). We used the version of CollaboRATE with a five-point response scale (28) and created a customised opening statement that asked participants to consider the conversation they had with health care providers about contraception when responding.

We also used existing, adapted, or new items to collect other data including age, clinic, ability to read and write English or Spanish, visit completion, previous study participation, sex assigned at birth (38), current gender identity (38), ethnicity (39), race (39), educational attainment (39), health insurance coverage (40), health literacy (41–45), reproductive history, and previous contraceptive method(s) (i.e., what, if any, contraceptive method(s) participants used in the four weeks before the health care visit). In addition, survey completion date, chosen survey language, and, where relevant, Internet Protocol (IP) address were recorded.

### Strategies for maximising enrolment, retention, and data quality

Informed by empirical evidence (46,47) and patient and stakeholder input, we adopted several strategies for achieving adequate participant enrolment, maximising participant retention, and enhancing the completeness and quality of data collected (see Appendix 8). Out of respect for people’s privacy, no attempts were made to ascertain why some eligible patients did not enrol in the study or were lost to follow-up.

### Sample size

In developing our study protocol, we performed detectable difference calculations for the primary outcome based on our anticipated capacity to enrol n=1,040 post-implementation cohort participants per trial arm during the study period and our assumption that 70% of these (i.e., n=728 post-implementation cohort participants per trial arm) would be included in the primary outcome analysis. These calculations are of lesser relevance due to changes in our analytic strategy (see *Analysis*) but are described in detail, along with relevant underlying assumptions, in our published study protocol (10).

### Analysis

In line with the pragmatic orientation of this trial (48), we analysed participants according to the trial arm of the clinic in which they received care, regardless of their exposure to the intervention(s). Analyses were conducted in SAS 9.4 and in SPSS Statistics v23-24.

#### Primary outcome

We assessed intervention effects on shared decision-making, scored as a binary variable, using a random effects logistic regression with a random intercept for clinic and adjustment for participant characteristics that differed across trial arms. We intended also to adjust for the clinic-level rate of shared decision-making during the pre-implementation data collection but because of the incompatibility of both adjusting for the clinic-level rate of shared decision-making and including a random intercept for clinic, we adopted an alternative strategy for reducing bias associated with non-equivalence of trial arms. Specifically, we included data collected from the pre-implementation cohort in the analytic model and, for each trial arm, calculated an odds ratio that represented the odds of shared decision-making among the post-implementation cohort compared to the pre-implementation cohort. We then conducted difference-in-differences analyses corresponding to our research questions and used the Scheffe Method to adjust for multiple comparisons.

We also examined heterogeneity of treatment effects on shared decision-making by assessing the significance of three-way interactions between cohort, trial arm, and each of several variables (age, health insurance coverage, health literacy, educational attainment, ethnicity and race, number of pregnancies, number of births, number of abortions, number of miscarriages, and previous contraceptive method(s)). We suggested possible variables for the heterogeneity of treatment effects analyses in our protocol (10) but the final variables and their coding were determined post-hoc, based on both theoretical reasons and subgroup sample sizes. We used p-value adjustment to account for the consequences of multiple comparisons. We followed up the significant three-way interaction identified by calculating intervention effect estimates separately among each subgroup.

#### Secondary outcomes

For secondary outcomes assessed at T1, we assessed intervention effects using the same methods as for the primary outcome. For the secondary outcomes assessed at T2 and T3, we adopted a different approach, deviating from our original plan. Specifically, we combined the data collected at T2 and at T3 in a single model, adding ‘time’ (i.e., T2 vs. T3) as a factor. This approach used all available data for participants who completed either or both of the follow-up surveys and thus represented an approach for handling missing data. For three secondary outcomes, we also conducted prespecified subgroup analyses to enhance the value of the study for diverse knowledge users (see Appendix 9).

#### Process outcomes

We calculated the proportion of participants in each trial arm who reported exposure to the relevant intervention(s). We also calculated the proportion of participants in each trial arm who reported exposure to the relevant intervention(s) that would recommend the intervention(s) to a friend.

#### Missing data

Handling of missing data varied across analyses and for some analyses deviated from our protocol (10). We excluded participants with missing data on the outcome of interest when we felt this was unlikely to undermine statistical power or produce biased estimates (i.e., where rates of missing data were very low, as for the outcomes and covariates assessed at T1). Where rates of missing data were higher (i.e., for outcomes assessed at T2 and T3), we used a different analytic approach to enable us to retain in analyses all participants who provided data on a particular outcome at T2 *or* T3, as described above (see *Secondary outcomes)*.

#### Planned analyses not conducted

We could not conduct analyses for five secondary outcomes (contraceptive method(s) used, use of a highly effective contraceptive method, unintended pregnancy (timing preferences), unintended pregnancy (pregnancy seeking), unwelcome pregnancy) and could not conduct one of the prespecified subgroup analyses for the secondary outcome of intended contraceptive method(s). In addition, we did not conduct planned comparisons pertaining to intervention feasibility or acceptability due to data reliability concerns.

#### Post hoc analyses

We chose to analyse the primary outcome as a binary variable based on its interpretability and its comparable psychometric properties to a continuous alternative in a previous study (28). However, we later questioned whether this choice had undermined statistical power to detect intervention effects and conducted post hoc analyses with shared decision-making scored as both a continuous variable and as a three-category variable. Additionally, after observing some heterogeneity of treatment effects on the primary outcome, we conducted post hoc analyses to explore whether there was heterogeneity of treatment effects on the secondary outcomes assessed at T1. Because these analyses identified very few substantive differences in intervention effect estimates among different subgroups, their results are not presented but are available on request.

### Patient and public involvement

The research team included three patient partners with different backgrounds and experiences (MB, PB, CCB), three stakeholder partners who were multidisciplinary health care providers with experience delivering contraceptive care in different settings and populations (MN, LFS, KKU), and one stakeholder partner with extensive experience in reproductive health advocacy (JN). We also held four 60-90 minute face-to-face listening sessions in three geographic regions at the outset of the project with patients diverse in age, ethnicity, race, and educational attainment. In these listening sessions, we solicited written and verbal feedback on proposed plans and draft materials and collaboratively generated study ideas and processes. We also formed a virtual Advisory Group of representatives from government and non-government organisations and efforts relevant to reproductive health care. The Advisory Group was engaged on four key occasions during the project and provided written feedback on proposed plans and draft materials. Patient and stakeholder input meaningfully impacted the design and execution of the study (see Appendix 10).

### Changes to study protocol

Refinements made to our analytic plan after trial commencement were described earlier (see *Analysis*) and other minor deviations from our published study protocol are described in Appendix 11.

### Ethical approval

Ethical approval for the study was granted by the Dartmouth College Committee for the Protection of Human Subjects (#00028721) and by the Care New England Memorial Hospital of Rhode Island Institutional Review Board (#855723-1).

## RESULTS

### Clinics

Participating clinics varied from very small, rural, community-based clinics to large, urban, hospital-based clinics. Twelve clinics were focused primarily on the provision of reproductive health care while four were focused primarily on the provision of primary care. Nine participating clinics were part of the Planned Parenthood network of health centres. The unadjusted rate of shared decision-making about contraceptive methods in each clinic among the entire pre-implementation cohort ranged from 34.5% to 72.2%. Where it could be estimated, the rate of patient participation in the study in each clinic varied from 11.6% to 70.5% (see Table 2).

**Table 2.** Characteristics of participating clinics by trial arm

|  | **Video**<br>(n=4) | **Decision aids**<br>(n=4) | **Dual**<br>(n=4) | **Usual care**<br>(n=4) |
| --- | --- | --- | --- | --- |
| **Primary focus of each clinic** |  |  |  |  |
| Reproductive health care | 3 | 3 | 4 | 2 |
| Primary care | 1 | 1 | 0 | 2 |
| **Unadjusted rate of shared decision-making about contraceptive methods among pre-implementation cohort (%)** |  |  |  |  |
| All clinics | 55.8 | 65.0 | 64.1 | 64.1 |
| Range among clinics | 34.5 – 67.9 | 48.7 – 71.3 | 48.4 – 72.2 | 37.5 – 70.7 |
| **Patient participation rate (%)<sup>1</sup>** |  |  |  |  |
| All clinics | 14.8 | 42.0 | 36.5 | 21.6 |
| Range at the clinic level | 11.6 – 37.3 | 30.3 – 70.5 | 32.4 – 42.8 | 17.4 – 29.0 |

*Notes.* <sup>1</sup>Calculated using the best available clinic-supplied data on the total number of unique eligible patients with health care visits during the study period. One clinic in the usual care arm was unable to supply relevant data and was excluded from these calculations.

### Participant flow

The flowchart of participants who consented to the study is presented in Figure 2. A flowchart of participants that provides data separately for the pre-implementation and post-implementation cohorts is provided in Appendix 12.

**Figure 2.**
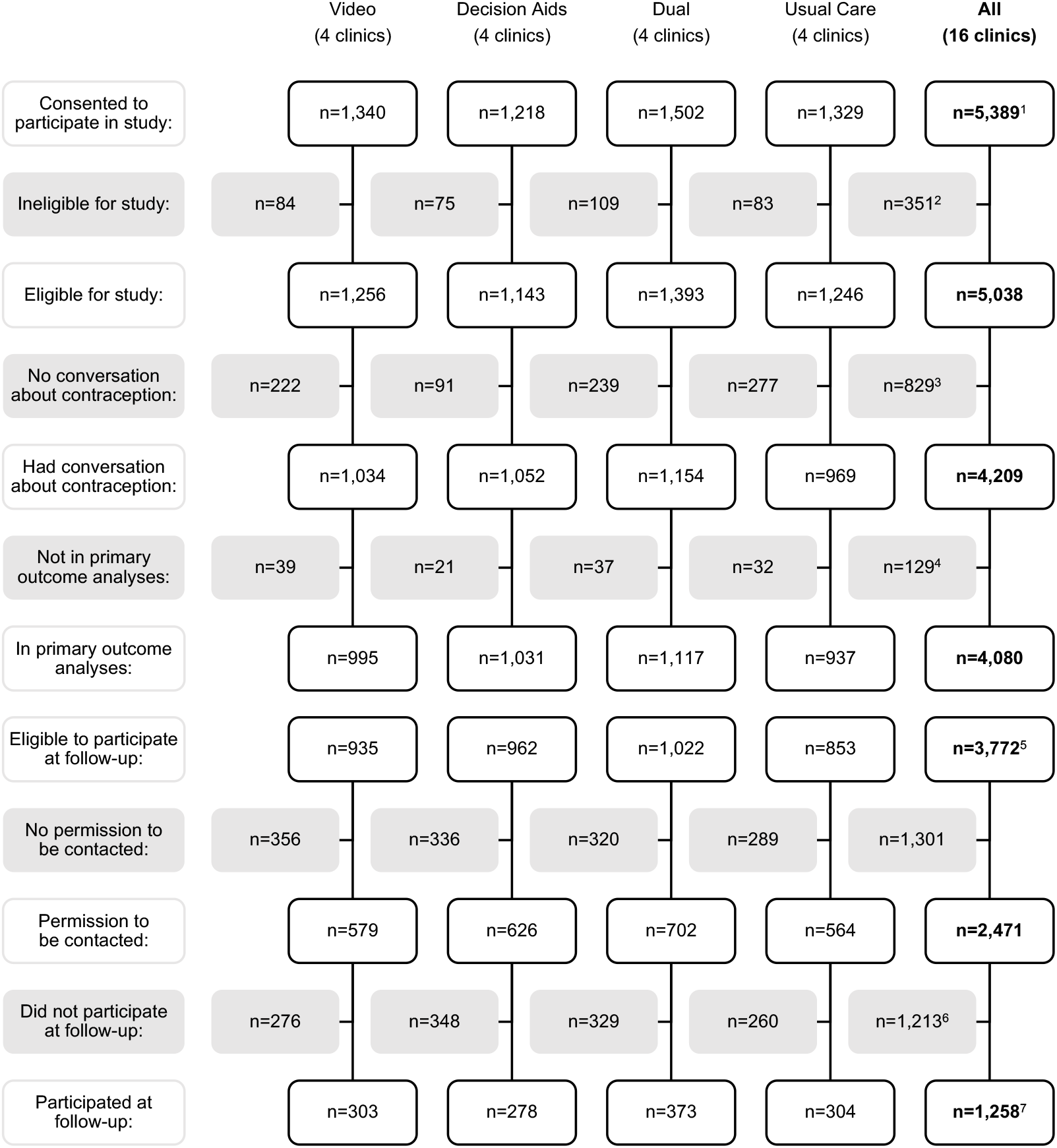
Flowchart of participants who consented to the study by trial arm ^1^A further 98 people who consented but had missing data on clinic name were excluded as their trial arm was unknown. ^2^Reasons were not reporting being able to read and write English or Spanish (n=10), not reporting no previous participation in the study (n=163), not reporting a completed health care visit (n=121), not reporting being aged 15-49 years (n=36), and not reporting being assigned female sex at birth (n=21). ^3^Reasons were not experiencing a conversation about contraception (n=809) and missing data on the item assessing occurrence of a conversation about contraception (n=20). ^4^Reasons were missing data on items assessing the primary outcome (n=21) and missing data on one or more covariates (n=108). ^5^Among those who experienced a conversation about contraception (n=4209), reasons for ineligibility were intending to use no contraceptive methods (n=411) and missing data on the item assessing intended contraceptive method(s) (n=26). ^6^Reasons were not being sent follow-up surveys due to discontinuing the T1 survey after giving permission to be contacted (n=6), returning surveys early or late (n=23), and other reasons, including incorrect, out of date, or missing contact details and electing not to participate (n=1184). ^7^287 participants completed only the T2 survey, 158 completed only the T3 survey, and 813 completed both the T2 and T3 surveys.

### Participants

The characteristics of participants who provided data on at least one study outcome are described by cohort and trial arm in Table 3.

**Table 3.**
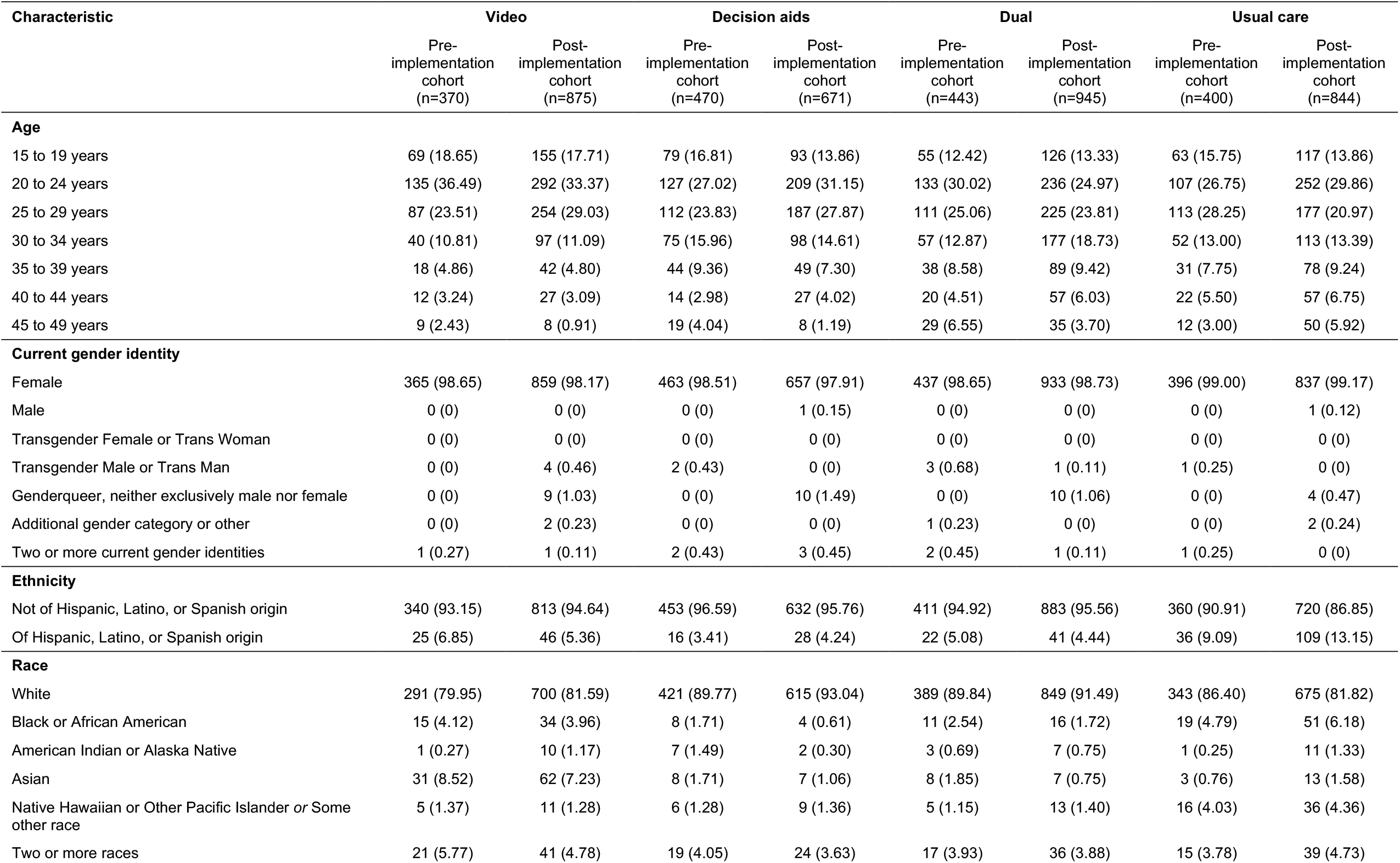
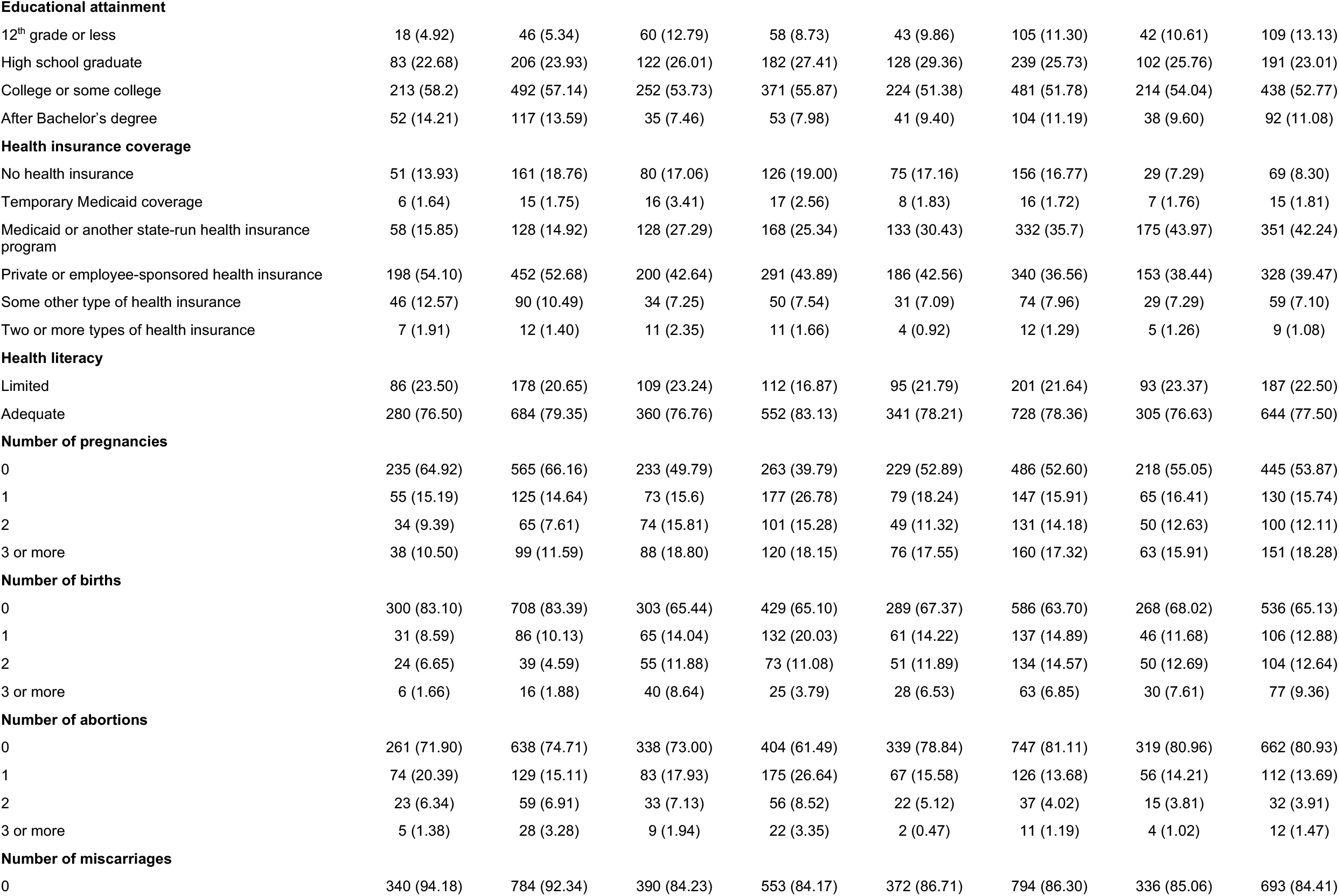
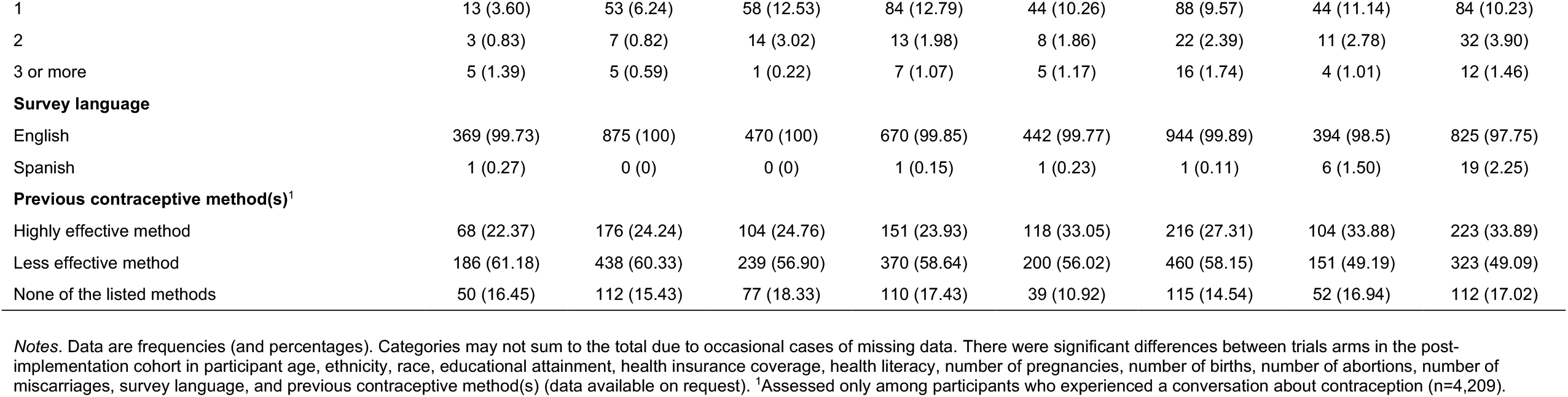
Characteristics of participants with data on at least one study outcome by trial arm and cohort (n=5,018)

| Characteristic | Video |  | Decision aids |  | Dual |  | Usual care |  |
| --- | --- | --- | --- | --- | --- | --- | --- | --- |
|  | Pre-implementation cohort (n=370) | Post-implementation cohort (n=875) | Pre-implementation cohort (n=470) | Post-implementation cohort (n=671) | Pre-implementation cohort (n=443) | Post-implementation cohort (n=945) | Pre-implementation cohort (n=400) | Post-implementation cohort (n=844) |
| **Age** |  |  |  |  |  |  |  |  |
| 15 to 19 years | 69 (18.65) | 155 (17.71) | 79 (16.81) | 93 (13.86) | 55 (12.42) | 126 (13.33) | 63 (15.75) | 117 (13.86) |
| 20 to 24 years | 135 (36.49) | 292 (33.37) | 127 (27.02) | 209 (31.15) | 133 (30.02) | 236 (24.97) | 107 (26.75) | 252 (29.86) |
| 25 to 29 years | 87 (23.51) | 254 (29.03) | 112 (23.83) | 187 (27.87) | 111 (25.06) | 225 (23.81) | 113 (28.25) | 177 (20.97) |
| 30 to 34 years | 40 (10.81) | 97 (11.09) | 75 (15.96) | 98 (14.61) | 57 (12.87) | 177 (18.73) | 52 (13.00) | 113 (13.39) |
| 35 to 39 years | 18 (4.86) | 42 (4.80) | 44 (9.36) | 49 (7.30) | 38 (8.58) | 89 (9.42) | 31 (7.75) | 78 (9.24) |
| 40 to 44 years | 12 (3.24) | 27 (3.09) | 14 (2.98) | 27 (4.02) | 20 (4.51) | 57 (6.03) | 22 (5.50) | 57 (6.75) |
| 45 to 49 years | 9 (2.43) | 8 (0.91) | 19 (4.04) | 8 (1.19) | 29 (6.55) | 35 (3.70) | 12 (3.00) | 50 (5.92) |
| **Current gender identity** |  |  |  |  |  |  |  |  |
| Female | 365 (98.65) | 859 (98.17) | 463 (98.51) | 657 (97.91) | 437 (98.65) | 933 (98.73) | 396 (99.00) | 837 (99.17) |
| Male | 0 (0) | 0 (0) | 0 (0) | 1 (0.15) | 0 (0) | 0 (0) | 0 (0) | 1 (0.12) |
| Transgender Female or Trans Woman | 0 (0) | 0 (0) | 0 (0) | 0 (0) | 0 (0) | 0 (0) | 0 (0) | 0 (0) |
| Transgender Male or Trans Man | 0 (0) | 4 (0.46) | 2 (0.43) | 0 (0) | 3 (0.68) | 1 (0.11) | 1 (0.25) | 0 (0) |
| Genderqueer, neither exclusively male nor female | 0 (0) | 9 (1.03) | 0 (0) | 10 (1.49) | 0 (0) | 10 (1.06) | 0 (0) | 4 (0.47) |
| Additional gender category or other | 0 (0) | 2 (0.23) | 0 (0) | 0 (0) | 1 (0.23) | 0 (0) | 0 (0) | 2 (0.24) |
| Two or more current gender identities | 1 (0.27) | 1 (0.11) | 2 (0.43) | 3 (0.45) | 2 (0.45) | 1 (0.11) | 1 (0.25) | 0 (0) |
| **Ethnicity** |  |  |  |  |  |  |  |  |
| Not of Hispanic, Latino, or Spanish origin | 340 (93.15) | 813 (94.64) | 453 (96.59) | 632 (95.76) | 411 (94.92) | 883 (95.56) | 360 (90.91) | 720 (86.85) |
| Of Hispanic, Latino, or Spanish origin | 25 (6.85) | 46 (5.36) | 16 (3.41) | 28 (4.24) | 22 (5.08) | 41 (4.44) | 36 (9.09) | 109 (13.15) |
| **Race** |  |  |  |  |  |  |  |  |
| White | 291 (79.95) | 700 (81.59) | 421 (89.77) | 615 (93.04) | 389 (89.84) | 849 (91.49) | 343 (86.40) | 675 (81.82) |
| Black or African American | 15 (4.12) | 34 (3.96) | 8 (1.71) | 4 (0.61) | 11 (2.54) | 16 (1.72) | 19 (4.79) | 51 (6.18) |
| American Indian or Alaska Native | 1 (0.27) | 10 (1.17) | 7 (1.49) | 2 (0.30) | 3 (0.69) | 7 (0.75) | 1 (0.25) | 11 (1.33) |
| Asian | 31 (8.52) | 62 (7.23) | 8 (1.71) | 7 (1.06) | 8 (1.85) | 7 (0.75) | 3 (0.76) | 13 (1.58) |
| Native Hawaiian or Other Pacific Islander or Some other race | 5 (1.37) | 11 (1.28) | 6 (1.28) | 9 (1.36) | 5 (1.15) | 13 (1.40) | 16 (4.03) | 36 (4.36) |
| Two or more races | 21 (5.77) | 41 (4.78) | 19 (4.05) | 24 (3.63) | 17 (3.93) | 36 (3.88) | 15 (3.78) | 39 (4.73) |
| **Educational attainment** |  |  |  |  |  |  |  |  |
| 12<sup>th</sup> grade or less | 18 (4.92) | 46 (5.34) | 60 (12.79) | 58 (8.73) | 43 (9.86) | 105 (11.30) | 42 (10.61) | 109 (13.13) |
| High school graduate | 83 (22.68) | 206 (23.93) | 122 (26.01) | 182 (27.41) | 128 (29.36) | 239 (25.73) | 102 (25.76) | 191 (23.01) |
| College or some college | 213 (58.2) | 492 (57.14) | 252 (53.73) | 371 (55.87) | 224 (51.38) | 481 (51.78) | 214 (54.04) | 438 (52.77) |
| After Bachelor's degree | 52 (14.21) | 117 (13.59) | 35 (7.46) | 53 (7.98) | 41 (9.40) | 104 (11.19) | 38 (9.60) | 92 (11.08) |
| **Health insurance coverage** |  |  |  |  |  |  |  |  |
| No health insurance | 51 (13.93) | 161 (18.76) | 80 (17.06) | 126 (19.00) | 75 (17.16) | 156 (16.77) | 29 (7.29) | 69 (8.30) |
| Temporary Medicaid coverage | 6 (1.64) | 15 (1.75) | 16 (3.41) | 17 (2.56) | 8 (1.83) | 16 (1.72) | 7 (1.76) | 15 (1.81) |
| Medicaid or another state-run health insurance program | 58 (15.85) | 128 (14.92) | 128 (27.29) | 168 (25.34) | 133 (30.43) | 332 (35.7) | 175 (43.97) | 351 (42.24) |
| Private or employee-sponsored health insurance | 198 (54.10) | 452 (52.68) | 200 (42.64) | 291 (43.89) | 186 (42.56) | 340 (36.56) | 153 (38.44) | 328 (39.47) |
| Some other type of health insurance | 46 (12.57) | 90 (10.49) | 34 (7.25) | 50 (7.54) | 31 (7.09) | 74 (7.96) | 29 (7.29) | 59 (7.10) |
| Two or more types of health insurance | 7 (1.91) | 12 (1.40) | 11 (2.35) | 11 (1.66) | 4 (0.92) | 12 (1.29) | 5 (1.26) | 9 (1.08) |
| **Health literacy** |  |  |  |  |  |  |  |  |
| Limited | 86 (23.50) | 178 (20.65) | 109 (23.24) | 112 (16.87) | 95 (21.79) | 201 (21.64) | 93 (23.37) | 187 (22.50) |
| Adequate | 280 (76.50) | 684 (79.35) | 360 (76.76) | 552 (83.13) | 341 (78.21) | 728 (78.36) | 305 (76.63) | 644 (77.50) |
| **Number of pregnancies** |  |  |  |  |  |  |  |  |
| 0 | 235 (64.92) | 565 (66.16) | 233 (49.79) | 263 (39.79) | 229 (52.89) | 486 (52.60) | 218 (55.05) | 445 (53.87) |
| 1 | 55 (15.19) | 125 (14.64) | 73 (15.6) | 177 (26.78) | 79 (18.24) | 147 (15.91) | 65 (16.41) | 130 (15.74) |
| 2 | 34 (9.39) | 65 (7.61) | 74 (15.81) | 101 (15.28) | 49 (11.32) | 131 (14.18) | 50 (12.63) | 100 (12.11) |
| 3 or more | 38 (10.50) | 99 (11.59) | 88 (18.80) | 120 (18.15) | 76 (17.55) | 160 (17.32) | 63 (15.91) | 151 (18.28) |
| **Number of births** |  |  |  |  |  |  |  |  |
| 0 | 300 (83.10) | 708 (83.39) | 303 (65.44) | 429 (65.10) | 289 (67.37) | 586 (63.70) | 268 (68.02) | 536 (65.13) |
| 1 | 31 (8.59) | 86 (10.13) | 65 (14.04) | 132 (20.03) | 61 (14.22) | 137 (14.89) | 46 (11.68) | 106 (12.88) |
| 2 | 24 (6.65) | 39 (4.59) | 55 (11.88) | 73 (11.08) | 51 (11.89) | 134 (14.57) | 50 (12.69) | 104 (12.64) |
| 3 or more | 6 (1.66) | 16 (1.88) | 40 (8.64) | 25 (3.79) | 28 (6.53) | 63 (6.85) | 30 (7.61) | 77 (9.36) |
| **Number of abortions** |  |  |  |  |  |  |  |  |
| 0 | 261 (71.90) | 638 (74.71) | 338 (73.00) | 404 (61.49) | 339 (78.84) | 747 (81.11) | 319 (80.96) | 662 (80.93) |
| 1 | 74 (20.39) | 129 (15.11) | 83 (17.93) | 175 (26.64) | 67 (15.58) | 126 (13.68) | 56 (14.21) | 112 (13.69) |
| 2 | 23 (6.34) | 59 (6.91) | 33 (7.13) | 56 (8.52) | 22 (5.12) | 37 (4.02) | 15 (3.81) | 32 (3.91) |
| 3 or more | 5 (1.38) | 28 (3.28) | 9 (1.94) | 22 (3.35) | 2 (0.47) | 11 (1.19) | 4 (1.02) | 12 (1.47) |
| **Number of miscarriages** |  |  |  |  |  |  |  |  |
| 0 | 340 (94.18) | 784 (92.34) | 390 (84.23) | 553 (84.17) | 372 (86.71) | 794 (86.30) | 336 (85.06) | 693 (84.41) |
| 1 | 13 (3.60) | 53 (6.24) | 58 (12.53) | 84 (12.79) | 44 (10.26) | 88 (9.57) | 44 (11.14) | 84 (10.23) |
| 2 | 3 (0.83) | 7 (0.82) | 14 (3.02) | 13 (1.98) | 8 (1.86) | 22 (2.39) | 11 (2.78) | 32 (3.90) |
| 3 or more | 5 (1.39) | 5 (0.59) | 1 (0.22) | 7 (1.07) | 5 (1.17) | 16 (1.74) | 4 (1.01) | 12 (1.46) |
| **Survey language** |  |  |  |  |  |  |  |  |
| English | 369 (99.73) | 875 (100) | 470 (100) | 670 (99.85) | 442 (99.77) | 944 (99.89) | 394 (98.5) | 825 (97.75) |
| Spanish | 1 (0.27) | 0 (0) | 0 (0) | 1 (0.15) | 1 (0.23) | 1 (0.11) | 6 (1.50) | 19 (2.25) |
| **Previous contraceptive method(s)<sup>1</sup>** |  |  |  |  |  |  |  |  |
| Highly effective method | 68 (22.37) | 176 (24.24) | 104 (24.76) | 151 (23.93) | 118 (33.05) | 216 (27.31) | 104 (33.88) | 223 (33.89) |
| Less effective method | 186 (61.18) | 438 (60.33) | 239 (56.90) | 370 (58.64) | 200 (56.02) | 460 (58.15) | 151 (49.19) | 323 (49.09) |
| None of the listed methods | 50 (16.45) | 112 (15.43) | 77 (18.33) | 110 (17.43) | 39 (10.92) | 115 (14.54) | 52 (16.94) | 112 (17.02) |

*Notes.* Data are frequencies (and percentages). Categories may not sum to the total due to occasional cases of missing data. There were significant differences between trials arms in the post-implementation cohort in participant age, ethnicity, race, educational attainment, health insurance coverage, health literacy, number of pregnancies, number of births, number of abortions, number of miscarriages, survey language, and previous contraceptive method(s) (data available on request). <sup>1</sup>Assessed only among participants who experienced a conversation about contraception (n=4,209).

### Shared decision-making about contraceptive methods

There was no difference in the odds of shared decision-making about contraceptive methods between pre- and post-implementation cohorts in any of the three intervention arms (see Table 4). In the usual care arm, the odds of shared decision-making about contraceptive methods were significantly lower in the post-implementation cohort than in the pre-implementation cohort (AOR=0.74, 95% CI: 0.55 to 0.98). However, difference-in-differences analyses corresponding to our research questions were non-significant. Post-hoc analyses conducted with shared decision-making scored as continuous and categorical variables yielded slightly different results (see Tables A6 and A7 in Appendix 13).

**Table 4.** Intervention effects on shared decision-making about contraceptive methods: differences between pre-implementation and post-implementation cohorts and difference-in-differences analyses

|  | Pre-implementation<br>cohort | Post-implementation<br>cohort | Difference between pre-implementation [REF] and<br>post-implementation<br>cohorts<br>AOR<sup>1</sup> (95% CI) |
| --- | --- | --- | --- |
|  | % | % |  |
| **All participants (n=4,080)** |  |  |  |
| Video (n=995) | 55.7 | 56.1 | 0.91 (0.68, 1.21) |
| Decision aids (n=1,031) | 65.7 | 69.2 | 1.08 (0.82, 1.42) |
| Dual (n=1,117) | 64.3 | 59.5 | 0.86 (0.65, 1.13) |
| Usual care (n=937) | 63.5 | 56.4 | **0.74 (0.55, 0.98)** |
| **Participants with adequate health literacy (n=3,234)** |  |  |  |
| Video (n=800) | 61.1 | 58.8 | 0.81 (0.58, 1.13) |
| Decision aids (n=828) | 70.8 | 70.7 | 0.98 (0.71, 1.34) |
| Dual (n=878) | 66.7 | 63.3 | 0.90 (0.66, 1.22) |
| Usual care (n=728) | 64.2 | 59.1 | 0.81 (0.58, 1.12) |
| **Participants with limited health literacy (n=846)** |  |  |  |
| Video (n=195) | 37.3 | 43.8 | 1.31 (0.70, 2.45) |
| Decision aids (n=203) | 50.0 | 61.4 | 1.44 (0.82, 2.55) |
| Dual (n=239) | 55.6 | 45.5 | 0.73 (0.42, 1.29) |
| Usual care (n=209) | 61.2 | 47.2 | **0.54 (0.30, 0.99)** |
|  | Difference-in-differences<br>AOR<sup>1</sup> (95% CI) | P-value unadjusted<br>for multiple<br>comparisons | P-value adjusted<br>for multiple<br>comparisons |
| **All participants (n=4,080)** |  |  |  |
| Video vs. Usual care [REF] | 1.23 (0.82, 1.85) | 0.32 | 0.80 |
| Decision aids vs. Usual care [REF] | 1.47 (0.98, 2.18) | 0.06 | 0.32 |
| Dual vs. Video [REF] | 0.95 (0.64, 1.41) | 0.79 | 1.00 |
| Dual vs. Decision aids [REF] | 0.80 (0.54, 1.17) | 0.25 | 0.72 |
| **Participants with adequate health literacy (n=3,234)** |  |  |  |
| Video vs. Usual care [REF] | 1.01 (0.63, 1.61) | 0.98 | 1.00 |
| Decision aids vs. Usual care [REF] | 1.21 (0.77, 1.91) | 0.41 | 0.88 |
| Dual vs. Video [REF] | 1.10 (0.70, 1.74) | 0.67 | 0.98 |
| Dual vs. Decision aids [REF] | 0.92 (0.59, 1.43) | 0.70 | 0.99 |
| **Participants with limited health literacy (n=846)** |  |  |  |
| Video vs. Usual care [REF] | **2.40 (1.01, 5.71)** | **0.05<sup>2</sup>** | 0.27 |
| Decision aids vs. Usual care [REF] | **2.65 (1.16, 6.07)** | **0.02** | 0.15 |
| Dual vs. Video [REF] | 0.56 (0.24, 1.30) | 0.18 | 0.61 |
| Dual vs. Decision aids [REF] | 0.51 (0.23, 1.13) | 0.10 | 0.43 |

Notes. AOR = Adjusted Odds Ratio, CI = Confidence Interval, REF = Reference group. <sup>1</sup>For all participants, analyses adjusted for age, health insurance coverage, health literacy, educational attainment, ethnicity, race, number of pregnancies, number of births, number of abortions, number of miscarriages, previous contraceptive method(s), and survey language. For participants with adequate health literacy and participants with limited health literacy, analyses adjusted for age, health insurance coverage, educational attainment, ethnicity, race, number of pregnancies, number of births, number of abortions, number of miscarriages, previous contraceptive method(s), and survey language. <sup>2</sup>p=0.047.

#### Heterogeneity of treatment effects on shared decision-making about contraceptive methods

There was a significant three-way interaction between trial arm, cohort (pre-implementation vs. post-implementation), and health literacy (adequate vs. limited) on shared decision-making about contraceptive methods (p<.0001) but all other interaction terms fitted for the heterogeneity of treatment effects analyses were not significant (see Table A8 in Appendix 13). To follow up the significant interaction between trial arm, cohort, and health literacy, we calculated intervention effect estimates separately for each health literacy subgroup (see Table 4).

Among participants with adequate health literacy, there was no difference in the odds of shared decision-making about contraceptive methods between pre- and post-implementation cohorts in any of the trial arms and difference-in-differences analyses corresponding to our research questions were non-significant.

Among participants with limited health literacy, there was no difference in the odds of shared decision-making about contraceptive methods between pre- and post-implementation cohorts in any of the three intervention arms. In the usual care arm, the odds of shared decision-making were significantly lower in the post-implementation cohort than in the pre-implementation cohort (AOR=0.54, 95% CI: 0.30 to 0.99). In addition, the difference between the pre- and post-implementation cohorts in the video arm was different from that in the usual care arm (AOR=2.40, 95% CI: 1.01 to 5.71) and the difference between the pre- and post-implementation cohorts in the decision aids arm was different from that in the usual care arm (AOR=2.65, 95% CI: 1.16 to 6.07). However, these differences were not robust to adjustment for multiple comparisons and other difference-in-differences analyses corresponding to our research questions were non-significant.

### Secondary outcomes

There were no differences between the pre- and post-implementation cohorts in any of the trial arms in the odds of having a conversation about contraception; the odds of satisfaction with the conversation about contraception; the odds of intending to use one or more contraceptive method(s); the odds of intending to use a highly effective contraceptive method; the odds of values concordance of the intended contraceptive method(s) immediately, four weeks, or six months after the health care visit; the odds of using the intended contraceptive method(s) four weeks after the health care visit; the odds of satisfaction with the contraceptive method(s) used four weeks after the health care visit; or mean decision regret about the intended contraceptive method(s) four weeks after the health care visit (see Table A9 in Appendix 14). Difference-in-differences analyses corresponding to our research questions for these outcomes were also non-significant (see Table A10 in Appendix 14). Where relevant, the same general findings emerged for the prespecified subgroup(s).

There were differences between the pre- and post-implementation cohorts in certain trial arms in the odds of adherence to the contraceptive method(s) used four weeks after the health care visit (decision aids arm: AOR=0.43, 95% CI: 0.19 to 0.97); the odds of adherence to the contraceptive method(s) used six months after the health care visit (decision aids arm: AOR=0.29, 95% CI: 0.10 to 0.84); the odds of satisfaction with the contraceptive method(s) used six months after the health care visit (dual interventions arm: AOR=0.32, 95% CI: 0.15 to 0.72); and mean decision regret about the intended contraceptive method(s) six months after the health care visit (decision aids arm: ALSM=0.21, 95% CI: 0.02 to 0.40) (see Table A9 in Appendix 14).

However, difference-in-differences analyses corresponding to our research questions for these outcomes were all non-significant after adjustment for multiple comparisons (see Table A10 in Appendix 14). In addition, although there was no difference between the pre- and post-implementation cohorts in any of the trial arms in the odds of using the intended contraceptive method(s) six months after the health care visit (see Table A9 in Appendix 14), the difference between the pre- and post-implementation cohorts in the decision aids arm was different from that in the usual care arm (AOR=0.45, 95% CI: 0.20 to 0.99; see Table A10 in Appendix 14). However, again, this difference was not robust to adjustment for multiple comparisons and other difference-in-differences analyses corresponding to our research questions were non-significant (see Table A10 in Appendix 14).

### Process outcomes

Among post-implementation cohort participants, 9.4% in the video arm said they watched the whole video and received the prompt card while waiting to see a health care provider, 31.5% in the decision aids arm said they used one of the decision aids together with a health care provider, and 5.0% in the dual interventions arm said they watched the whole video, received the prompt card, and used one of the decision aids (see Table A11 in Appendix 15). Notably, however, comparable data provided by pre-implementation cohort participants suggested some measurement error (e.g., see Table A11 in Appendix 15). We therefore recommend caution when interpreting rates of intervention implementation and drawing conclusions about intervention feasibility.

Among post-implementation cohort participants who said they watched the whole video while waiting to see a health care provider, 94.7% said they would recommend it to a friend. Among post-implementation cohort participants who said they received the prompt card while waiting to see a health care provider, 91.4% said they would recommend it to a friend. Among post-implementation cohort participants who said they used one of the decision aids together with a health care provider, 98.5% said they would recommend them to a friend (see Table A12 in Appendix 15).

## DISCUSSION

For the primary outcome of shared decision-making about contraceptive methods, we observed no difference between the pre- and post-implementation cohorts in any of the three intervention arms and found no difference-in-differences when making comparisons between relevant arms. For participants with adequate health literacy, results largely resembled those for all participants. However, there were slightly different results for those with limited health literacy. First, there was a pronounced difference in shared decision-making between the pre- and post-implementation cohorts in the usual care arm. Second, the difference in shared decision-making between the pre- and post-implementation cohorts was different in the video arm and in the decision aids arm from that in the usual care arm. These differences were not robust to adjustment for multiple comparisons. However, we consider this more likely due to inadequate power to detect intervention effects in this subgroup than to a true absence of them. Indeed, clinic staff members we interviewed perceived that the interventions studied were particularly valuable for patients with lower literacy and those making health care decisions independently for the first time (26).

The absence of intervention effects on shared decision-making in the dual interventions arm was unexpected. A systematic review of interventions for improving adoption of shared decision-making cautiously concluded that interventions targeting both patients and providers were generally more effective than those targeting either group alone (6). However, it could be that such findings occur only under ideal conditions with high implementation fidelity. Although interviews with clinic staff members suggested that those from the dual interventions arm did not perceive a substantially higher implementation burden than those from single intervention arms (26), there may have been lower fidelity of intervention implementation in the dual interventions arm.

The observed difference in shared decision-making about contraceptive methods between the pre-implementation and post-implementation cohorts in the usual care arm was also unexpected. One explanation for this finding is that introduction of measurement of shared decision-making in clinics, which was not concealed from clinic staff, had an initial but unsustained positive effect on the behaviour (49). A second is that the pre-implementation cohort was systematically different from the post-implementation cohort on some unmeasured variable(s) correlated with shared decision-making. A third is that changing political or other environmental factors (e.g., staff turnover, changes in provider workload or priorities) accounted for the observed decrease. Regardless of the explanation, however, we have no reason to believe that our intervention arms would have been impervious to the factors responsible for the decrease in the usual care arm, which was important for our interpretation of findings.

### Limitations and strengths

There were some limitations. Conducting this study in only the Northeast United States may have caused us to underrepresent some populations, including people of colour, and may have impacted the generalisability of findings. The number of clinics also prevented stratified assignment and heterogeneity of treatment effect analyses based on clinic type. Rates of participant retention were lower than anticipated and therefore results for secondary outcomes assessed at follow-up are vulnerable to nonresponse bias. In addition, we were unable to confidently describe fidelity of implementation of the various intervention components. However, these limitations are balanced by several strengths. Enrolling the pre-implementation cohort enabled us to adopt stratified assignment of clinics to trial arms based on the initial rate of shared decision-making about contraceptive methods, to observe the decrease in shared decision-making in the usual care arm (and thus more accurately interpret patterns in the intervention arms), to conduct difference-in-differences analyses, and to be alert to possible measurement error in patient-reported data on intervention exposure. Our highly pragmatic trial – which included small, rural, and community-based clinics and participants across the reproductive age range, allowed the use of existing or new concomitant interventions and care, and evaluated interventions not reliant on resources or infrastructure not usually available in real-world settings – resulted in new insights into whether patient decision aids and question prompt interventions are likely to meaningfully impact shared decision-making in practice (48). That under pragmatic conditions there may have been only modest intervention uptake and were few intervention effects is very important, especially given widespread advocacy of these kinds of interventions without due attention to issues of feasibility and real-world impact.

### Recommendations for future research

This work has exposed several areas of uncertainty that represent valuable research directions. Most importantly, we recommend greater attention be dedicated to understanding strategies that encourage and enable the uptake of efficacious shared decision-making interventions under real-world conditions. Although the ‘light-touch’ approaches we used may have been inadequate, there may well be other scalable strategies that foster the intervention uptake necessary for meaningful improvements in patient experiences and outcomes. We also recommend that research seek to understand the decrease in shared decision-making we observed in the usual care arm. If the determinants of this finding are not specific to our study context, there are implications for other research, particularly for comparative effectiveness studies that do not employ a usual care arm and thus cannot capture such changes. Finally, we recommend that research be dedicated to understanding how to reliability assess fidelity in intervention delivery in large-scale research studies where observational methods are not feasible or would function as an additional intervention. Specific questions include how to generate patient-reported data on intervention exposure that is not vulnerable to measurement error (whether due to confusion, uncertainty, or social desirability), and how to efficiently triangulate such data with data from other sources.

## Conclusions

In this study, we observed no effects of the video and prompt card or the decision aids and training on shared decision-making about contraceptive methods whether implemented alone or together. We conclude that these interventions are unlikely to have a meaningful population-wide impact on shared decision-making in real-world contraceptive care without additional strategies to encourage and enable implementation. Our interventions appeared more promising for patients with limited health literacy and, if effective, could reduce important disparities in shared decision-making. However, further evaluation in a trial powered to detect intervention effects in this subgroup is needed, as is consideration of whether selective implementation of the interventions would prove more or less feasible than the universal implementation attempted in this study.

## Supporting information

Reporting Checklist

## Data Availability

Requests for access to anonymised data for research purposes may be submitted to the corresponding author.

## Disclaimer

The statements, findings, and conclusions presented in this paper are solely the responsibility of the authors and do not necessarily represent the views of the Patient-Centered Outcomes Research Institute® (PCORI®), its Board of Governors or Methodology Committee, the National Institutes of Health, or the Planned Parenthood Federation of America, Inc.

## Transparency statement

The lead author affirms that the manuscript is an honest, accurate, and transparent account of the study being reported; that no important aspects of the study have been omitted; and that any discrepancies from the study as originally planned (and, if relevant, registered) have been explained.

## Contributions

RT, KZD, and GE conceived the study. RT, GS, and RM led development and implementation of the data collection strategy, including survey design and development, patient-reported measure development, survey and survey distribution protocol development and programming, and data management. KD led liaison with participating clinics and clinic representatives. DA led adaptation of interventions, patient-reported measures, surveys, and other study materials for Spanish-speaking patients. ZL and TDT led development of the analytic plan and conducted the inferential analyses. All authors contributed to the design of the study (e.g., selection of interventions, development of participant and clinic eligibility criteria, selection of assignment approach, selection of study outcomes); the development of study interventions, recruitment materials (e.g., study posters, information sheets) and data collection materials and protocols (e.g., patient surveys, survey invitation and reminder schedules); and/or the interpretation of data. RT and GS drafted the manuscript content. All authors contributed to revising the draft manuscript content and gave final approval of the version to be published. All authors agree to be accountable for all aspects of the work in ensuring that questions related to the accuracy or integrity of any part of the work are appropriately investigated and resolved.

## Acknowledgments

We are grateful to Amina Hetu and Nitzy Bustamante for their contribution to the development of study materials, to Shama Alam and Elizabeth Harman for participating in the production of study interventions, and to those who provided valuable feedback on early drafts of study materials, including Amanda Beery, Rachel Darche, Ann Davis, Amanda Dennis, Candace Gibson, Christina Lachance, Lindsay Smith, Michele Stranger-Hunter, Lawrence Swiader, Christina Warner, Jacki Witt, Elisabeth Woodhams, and Lauren Zapata.

