## Appendices for "Right For Me: a pragmatic multi-arm cluster randomised controlled trial of two interventions for increasing shared decision-making about contraceptive methods"


| | |
|---|---|
| **APPENDIX 1** | Research questions and hypotheses |
| **APPENDIX 2** | Study poster and information sheet |
| **APPENDIX 3** | Right For Me prompt card |
| **APPENDIX 4** | Right For Me decision aids |
| **APPENDIX 5** | Right For Me written guidance |
| **APPENDIX 6** | Implementation slide deck: Video + Prompt Card |
| **APPENDIX 7** | Implementation slide deck: Decision Aids + Training |
| **APPENDIX 8** | Strategies for maximising enrolment, retention, and data quality |
| **APPENDIX 9** | Outcomes and measures |
| **APPENDIX 10** | Impact of patient and public involvement |
| **APPENDIX 11** | Protocol refinements and deviations |
| **APPENDIX 12** | Participant flow |
| **APPENDIX 13** | Supplementary results tables: primary outcome |
| **APPENDIX 14** | Supplementary results tables: secondary outcomes |
| **APPENDIX 15** | Supplementary results tables: process outcomes |

#### APPENDIX 1 Research questions and hypotheses

**Table A1.** Research questions and hypotheses

| Research Questions | Hypotheses |
|---|---|
| **OBJECTIVE 1 (PRIMARY OUTCOME)** | |
| 1. Does implementing the video + prompt card increase the rate of shared decision-making about contraceptive methods compared to usual care? | Implementing the video + prompt card will increase the rate of shared decision-making about contraceptive methods compared to usual care. |
| 2. Does implementing the decision aids + training increase the rate of shared decision-making about contraceptive methods compared to usual care? | Implementing the decision aids + training will increase the rate of shared decision-making about contraceptive methods compared to usual care. |
| 3. Does implementing the video + prompt card and the decision aids + training result in greater increases in the rate of shared decision-making about contraceptive methods compared to usual care than implementing either of the interventions alone? | Implementing the video + prompt card and the decision aids + training will result in greater increases in the rate of shared decision-making about contraceptive methods compared to usual care than implementing the video + prompt card alone or the decision aids + training alone. |
| 4. What patient characteristics and other factors modify the effect of implementing the interventions on the rate of shared decision-making about contraceptive methods? | This heterogeneity of treatment effects analysis is exploratory (i.e., hypothesis generating) and thus no a priori hypotheses have been developed. |
| **OBJECTIVE 2 (SECONDARY OUTCOMES)** | |
| *For each of the secondary outcomes:* | |
| 5. Does implementing the video + prompt card increase or decrease (as relevant) the [rate/level] of [secondary outcome] compared to usual care? | Implementing the video + prompt card will increase the rate of conversation about contraception, satisfaction with the conversation about contraception, values concordance of intended contraceptive method(s), use of intended contraceptive method(s), adherence to contraceptive method(s) used, and satisfaction with contraceptive method(s) used; and decrease the level of decision regret, the rate of unintended pregnancy[1], and the rate of unwelcome pregnancy compared to usual care. Analyses pertaining to intended contraceptive method(s), intention to use a highly effective contraceptive method, contraceptive method(s) used, and use of a highly effective contraceptive method are exploratory and thus no a priori hypotheses have been developed. |
| 6. Does implementing the decision aids + training increase or decrease (as relevant) the [rate/level] of [secondary outcome] compared to usual care? | Implementing the decision aids + training will increase the rate of conversation about contraception, satisfaction with the conversation about contraception, values concordance of intended contraceptive method(s), use of intended contraceptive method(s), adherence to contraceptive method(s) used, and satisfaction with contraceptive method(s) used; and decrease the level of decision regret, the rate of unintended pregnancy[1], and the rate of unwelcome pregnancy compared to usual care. Analyses pertaining to intended contraceptive method(s), intention to use a highly effective contraceptive method, contraceptive method(s) used, and use of a highly effective contraceptive method are exploratory and thus no a priori hypotheses have been developed. |
| 7. Does implementing the video + prompt card and the decision aids + training result in greater increases or decreases (as relevant) in the [rate/level] of [secondary outcome] compared to usual care than implementing either of the interventions alone? | Implementing the video + prompt card and the decision aids + training will result in greater increases in the rate of conversation about contraception, satisfaction with the conversation about contraception, values concordance of intended contraceptive method(s), use of intended contraceptive method(s), adherence to contraceptive method(s) used, and satisfaction with contraceptive method(s) used; and greater decreases in the level of decision regret, the rate of unintended pregnancy[1], and the rate of unwelcome pregnancy compared to usual care than implementing the video + prompt card alone or the decision aids + training alone. Analyses pertaining to intended contraceptive method(s), intention to use a highly effective contraceptive method, contraceptive method(s) used, and use of a highly effective contraceptive method are exploratory and thus no a priori hypotheses have been developed. |
| **OBJECTIVE 3 (PROCESS OUTCOMES)** | |
| 8. Of participants receiving care in a trial arm implementing the video + prompt card, what proportion report having watched the whole video? | Of participants receiving care in a trial arm implementing the video + prompt card, at least 70% will report having watched the whole video. |
| 9. Of participants receiving care in a trial arm implementing the video + prompt card, what proportion report having received the prompt card? | Of participants receiving care in a trial arm implementing the video + prompt card, at least 70% will report having received the prompt card. |

| | |
|---|---|
| 10. Of participants receiving care in a trial arm implementing the decision aids + training, what proportion report having used a decision aid together with a health care provider? | Of participants receiving care in a trial arm implementing the decision aids + training, at least 70% will report having used a decision aid together with a health care provider. |
| 11. Is the proportion of participants who report having watched the whole video higher among those receiving care in a trial arm implementing the video + prompt card and the decision aids + training than in a trial arm implementing the video + prompt card alone? | The proportion of participants who report having watched the whole video will be higher among those receiving care in a trial arm implementing the video + prompt card and the decision aids + training than in a trial arm implementing the video + prompt card alone. |
| 12. Is the proportion of participants who report having received the prompt card higher among those receiving care in a trial arm implementing the video + prompt card and the decision aids + training than in a trial arm implementing the video + prompt card alone? | The proportion of participants who report having received the prompt card will be higher among those receiving care in a trial arm implementing the video + prompt card and the decision aids + training than in a trial arm implementing the video + prompt card alone. |
| 13. Is the proportion of participants who report having used a decision aid together with a health care provider higher among those receiving care in a trial arm implementing the video + prompt card and the decision aids + training than in a trial arm implementing the decision aids + training alone? | The proportion of participants who report having used a decision aid together with a health care provider will be higher among those receiving care in a trial arm implementing the video + prompt card and the decision aids + training than in a trial arm implementing the decision aids + training alone. |
| 14. What proportion of participants who report having watched the whole video would recommend it to a friend? | A majority of participants who report having watched the whole video would recommend it to a friend. |
| 15. What proportion of participants who report having received the prompt card would recommend it to a friend? | A majority of participants who report having received the prompt card would recommend it to a friend. |
| 16. What proportion of participants who report having used a decision aid together with a health care provider would recommend it to a friend? | A majority of participants who report having used a decision aid together with a health care provider would recommend it to a friend. |
| 17. Is the proportion of participants who would recommend the video to a friend higher among those receiving care in a trial arm implementing the video + prompt card and the decision aids + training than in a trial arm implementing the video + prompt card alone? | The proportion of participants who would recommend the video to a friend will be higher among those receiving care in a trial arm implementing the video + prompt card and the decision aids + training than in a trial arm implementing the video + prompt card alone. |
| 18. Is the proportion of participants who would recommend the prompt card to a friend higher among those receiving care in a trial arm implementing the video + prompt card and the decision aids + training than in a trial arm implementing the video + prompt card alone? | The proportion of participants who would recommend the prompt card to a friend will be higher among those receiving care in a trial arm implementing the video + prompt card and the decision aids + training than in a trial arm implementing the video + prompt card alone. |
| 19. Is the proportion of participants who would recommend the decision aids to a friend higher among those receiving care in a trial arm implementing the video + prompt card and the decision aids + training than in a trial arm implementing the decision aids + training alone? | The proportion of participants who would recommend the decision aids to a friend will be higher among those receiving care in a trial arm implementing the video + prompt card and the decision aids + training than in a trial arm implementing the decision aids + training alone. |

*Notes.* [1]Unintended pregnancy was measured based both on pregnancy timing preferences immediately before conception and pregnancy seeking immediately before conception.

**APPENDIX 2** Study poster and information sheet

right for me

This clinic is involved in the Right For Me study
Participating involves doing a brief health care survey

#### You can participate if you:

had a **health care visit** at this clinic today

can read and write **English** or **Spanish**

are **15 to 49** years old

were assigned **female** sex at birth

have **not participated before**

#### Participants receive a $10 gift card

To participate, please ask a clinic staff member

You can participate in this study if you:

had a **health care visit** at this clinic today

can read and write **English** or **Spanish**

are **15 to 49** years old

were assigned **female** sex at birth

have **not participated before**

We will check that you meet these criteria at the start of the study

#### Who can I contact?

If you have questions or concerns about this study, you can contact:


**Ruth Manski**
Right For Me Project Manager
Dartmouth College
(e)
(t) 603 653 0879
(during business hours)

**Rachel Thompson**
Right For Me Principal Investigator
Dartmouth College
(e)
(t) 603 653 0860
(during business hours)


If you have questions, concerns, complaints, or suggestions about research at Dartmouth, you can call:

**Office of the Committee for the Protection of Human Subjects**
Dartmouth College
(t) 603 646 6482
(during business hours)

THE
Dartmouth
INSTITUTE
FOR HEALTH POLICY & CLINICAL PRACTICE


This study is funded through a Patient-Centered Outcomes Research Institute (PCORI) Award (CDR-1403-12221).


right for me

### Study Information

Right For Me is a research study led by Dartmouth College. Your clinic is involved in the study.

This handout gives information about the Right For Me study.

If there's something you don't understand, please ask questions.

#### What is the purpose?

The purpose of this study is to learn new things about people's experiences of health care and how improvements can be made to help people get the health care they want and need.

#### What is involved?

Participating involves doing a 5-minute survey on a tablet computer before you leave the clinic. The survey asks about your health care today. If relevant, the survey also asks about your experiences and views about birth control.

A small group of people will be invited to do two more 5-minute surveys at home later. The surveys ask more questions about experiences and views about birth control. The first survey will be sent in 4 weeks. The second survey will be sent in 6 months.

#### Do I have to participate?

No, participating is voluntary. Your choice about whether to participate will not affect the quality of your health care. You can also stop participating at any time. If you stop participating, the information we have already collected will continue to be used.

#### How many people will participate?

This is a large and important study. Altogether, 16 clinics from New England are involved. We expect that 390 people from your clinic and 5,850 people from other clinics will participate.

#### Will I be paid?

You will receive a $10 gift card for Amazon.com for every survey you complete. If you are aged under 20, you will need an email address to receive the gift card. If you are aged 20 or older, you will need either an email address or a mailing address. The gift card will be sent no more than two weeks after you complete each survey.

If the information we collect in the study is used to develop a product sold for a profit, you will not receive a share of the profits.

#### What are the benefits?

You may enjoy completing the survey and find it valuable to give feedback on your health care. People receiving health care in the future may also benefit from what we learn from the study.

#### What are the risks?

There are risks involved in most life activities. We think that the risks involved in participating in this study are minimal. You may find some of the survey questions personal. You can skip any questions you don't want to answer and remain in the study.

#### How will you use the information?

The information we collect will be stored for an indefinite period of time.

We will use the information we collect to learn new things about people's experiences of health care and how improvements can be made. We will share what we learn with participants, researchers, health professionals, people who design health care, and the public.

We may also share the information we collect with other people so that they can confirm what we learned or do new studies on related topics.

#### How will you protect my privacy?

Protecting your privacy is important to us.

Some of the information we collect could identify you. Information that could identify you will be treated as confidential.

None of the people that provide health care in clinics during the study will be allowed to see information that could identify you.

The people that will be allowed to see information that could identify you are: (1) the study principal investigator, project manager, and data manager, and (2) people who work in information security, information management, and translation at Dartmouth College and elsewhere. It is also possible for a court or government official to order the release of study data including information about you.

If you choose to receive your gift card via email, we will give your email address to a company so they can email it to you.

We will store information that could identify you in password-protected computer files and locked filing cabinets. We will only ever transfer this information using secure methods.

We will remove or change all information that could identify you before we share what we learn with others. We will also remove or change all information that could identify you before we share the information we collect with others.

#### How can I find out what you learn?

You can find out what we learn from the study by visiting the study website (www.rightforme.org) after January 2018.

**APPENDIX 3** Right For Me prompt card

### Try asking these questions today

What are my options?

What are the possible pros and cons of those options?

How likely are each of those pros and cons to happen to me?

right for me

**APPENDIX 4** Right For Me decision aids

### Types of Birth Control Methods

This decision aid is to help you and your health care provider talk about methods of birth control and choose what's right for you.
Most people can safely use these methods. Your health care provider can tell you whether these methods are safe for you.

right for me

| | LONG-ACTING | SHORT-ACTING | BARRIER | NATURAL* | PERMANENT | EMERGENCY |
|---|---|---|---|---|---|---|
| What are they? | Birth control methods that are placed by a health care provider and last between 3 and 10 years | Birth control methods that are used every day, every week, every 4 weeks, or every 13 weeks | Birth control methods that are used every time you have sex | Birth control methods that do not involve any hormones or devices | Birth control methods that involve a procedure to close off the tubes that carry eggs or sperm | Birth control methods that are used after unprotected sex |
| What are the options? | • Implant<br>• Hormonal IUD<br>• Copper IUD | • Injection<br>• Progestin Pill<br>• Combined Pill<br>• Patch<br>• Ring | • Male Condom<br>• Female Condom<br>• Spermicide<br>• Sponge<br>• Cervical Cap<br>• Diaphragm | • Withdrawal Method<br><br>*Fertility Awareness Methods:*<br>• Standard Days Method®<br>• TwoDay Method®<br>• Ovulation Method<br>• Symptothermal Method | *Female Sterilization:*<br>• By Laparoscopy<br>• By Minilaparotomy<br>• By Hysteroscopy<br><br>*Male Sterilization:*<br>• Vasectomy | • Copper IUD<br>• Ulipristal Pill<br>• Progestin Emergency Pill<br>• Combined Pill |
| Who might choose them? | People who want or are comfortable with:<br><br>• Almost no chance of pregnancy (fewer than 1 in 100 people become pregnant in the first year)<br><br>• A method they can almost forget about<br><br>• A procedure to start and stop using the method | People who want or are comfortable with:<br><br>• Some chance of pregnancy (6 to 9 in 100 people become pregnant in the first year)<br><br>• A method they need to remember<br><br>• A hormonal method<br><br>• A method they can stop without a health care visit | People who want or are comfortable with:<br><br>• A higher chance of pregnancy (12 to 29 in 100 people become pregnant in the first year)<br><br>• A method they need to remember<br><br>• A non-hormonal method<br><br>• Protection against sexually transmitted infections (STIs) (not all methods offer this) | People who want or are comfortable with:<br><br>• A higher chance of pregnancy (22 to 24 in 100 people become pregnant in the first year)<br><br>• A method they need to remember<br><br>• A non-hormonal method<br><br>• A method that does not involve birth control devices | People who want or are comfortable with:<br><br>• Almost no chance of pregnancy (fewer than 1 in 100 people become pregnant in the first year)<br><br>• A method they can forget about<br><br>• A procedure<br><br>• Never becoming pregnant in the future | People who:<br><br>• Have had unprotected sex and don't want to become pregnant |

*The Lactational Amenorrhea Method is another natural birth control method that may be used by some people who are breastfeeding. Your health care provider can tell you about this method.



© 2016 Trustees of Dartmouth College | Version 1.0

### Long-Acting Reversible Birth Control Methods

This decision aid is to help you and your health care provider talk about methods of birth control and choose what's right for you.
Most people can safely use these methods. Your health care provider can tell you whether these methods are safe for you.

right for me

| | IMPLANT | HORMONAL IUD | COPPER IUD |
|---|---|---|---|
| **How is it used?** | A health care provider puts a flexible device the size of a matchstick under the skin of your arm | A health care provider puts a small, T-shaped, plastic device in your uterus | A health care provider puts a small, T-shaped plastic and copper device in your uterus |
| **How often?** | Every 3 years | Every 3 to 5 years<br>(depends on the brand) | Every 10 years |
| **How does it work?** | Releases progestin | Releases progestin | Releases copper |
| **When does it start working?** | Immediately or after 7 days<br>(depends on when you get the implant) | Immediately or after 7 days<br>(depends on when you get the IUD) | Immediately |
| **How many people become pregnant in the first year?** | | | |
| Not always following the instructions: | Fewer than 1 in 100 people<br>○○○○○○○○○○ | Fewer than 1 in 100 people<br>○○○○○○○○○○ | Fewer than 1 in 100 people<br>○○○○○○○○○○ |
| Always following the instructions: | Fewer than 1 in 100 people<br>○○○○○○○○○○ | Fewer than 1 in 100 people<br>○○○○○○○○○○ | Fewer than 1 in 100 people<br>○○○○○○○○○○ |
| **What are some of the side effects?** | | | |
| Unscheduled spotting or bleeding? | Possible, may or may not improve over time | Possible, usually improves over time | Possible, usually improves over time |
| Heavy or prolonged bleeding? | Possible | | Possible, usually improves over time |
| More painful periods? | | | Possible, usually improves over time |
| Fewer or no periods? | Possible | Possible | |
| Other side effects (e.g., nausea, headaches, breast tenderness, mood changes or depression)? | Possible | Possible | |
| Device comes out? | | Possible | Possible |
| **Does it protect against sexually transmitted infections (STIs)?** | No | No | No |

For more information, including authors, information sources, and terms of use, see www.rightforme.org/decision

© 2016 Trustees of Dartmouth College | Version 1.0

### Short-Acting Reversible Birth Control Methods

right for me

This decision aid is to help you and your health care provider talk about methods of birth control and choose what's right for you. Most people can safely use these methods. Your health care provider can tell you whether these methods are safe for you.

| | INJECTION | PROGESTIN PILL | COMBINED PILL | PATCH | RING |
|---|---|---|---|---|---|
| **How is it used?** | A health care provider gives you a shot in your arm or buttock | You swallow a small pill | You swallow a small pill | You stick a small, thin patch on your body | You put a flexible, plastic ring in your vagina |
| **How often?** | Every 13 weeks | Every day at the same time | Every day at the same time | Every week | Every 4 weeks |
| **How does it work?** | Releases progestin | Releases progestin | Releases progestin and estrogen | Releases progestin and estrogen | Releases progestin and estrogen |
| **When does it start working?** | Immediately or after 7 days (depends on when you get the first shot) | Immediately or after 2 days (depends on when you start taking the pill) | Immediately or after 7 days (depends on when you start taking the pill) | Immediately or after 7 days (depends on when you start using the patch) | Immediately or after 7 days (depends on when you start using the ring) |
| **How many people become pregnant in the first year?** | | | | | |
| Not always following the instructions: | 6 in 100 people ●○○○○○○○○○ | 9 in 100 people ●○○○○○○○○○ | 9 in 100 people ●○○○○○○○○○ | 9 in 100 people ●○○○○○○○○○ | 9 in 100 people ●○○○○○○○○○ |
| Always following the instructions: | Fewer than 1 in 100 people ○○○○○○○○○○ | Fewer than 1 in 100 people ○○○○○○○○○○ | Fewer than 1 in 100 people ○○○○○○○○○○ | Fewer than 1 in 100 people ○○○○○○○○○○ | Fewer than 1 in 100 people ○○○○○○○○○○ |
| **What are some of the side effects?** | | | | | |
| Unscheduled spotting or bleeding? | Possible | Possible | Possible, usually improves over time | Possible, usually improves over time | Possible, usually improves over time |
| Heavy or prolonged bleeding? | Possible | Possible | | | |
| Fewer or no periods? | Possible | Possible | | | |
| Other side effects (e.g., nausea, headaches, breast tenderness)? | | Possible | Possible | Possible | Possible |
| Bone loss? | Possible, usually improves when you stop getting the shot | | | | |
| Skin irritation? | | | | Possible | |
| Vaginal irritation or discharge? | | | | | Possible |
| **Does it protect against sexually transmitted infections (STIs)?** | No | No | No | No | No |



© 2016 Trustees of Dartmouth College | Version 1.0

right for me

### Barrier Birth Control Methods

This decision aid is to help you and your health care provider talk about methods of birth control and choose what's right for you. Most people can safely use these methods. Your health care provider can tell you whether these methods are safe for you.

| | MALE CONDOM | FEMALE CONDOM | SPERMICIDE | SPONGE | CERVICAL CAP (with spermicide) | DIAPHRAGM (with spermicide) |
|---|---|---|---|---|---|---|
| **How is it used?** | A thin latex or polyurethane sleeve is put on the erect penis before sex | You put a thin sleeve in your vagina before sex | You put a foam, cream, gel, film, or suppository in your vagina before sex | You put a round foam device (that already contains spermicide) in your vagina before sex | You put a silicone-rubber device in your vagina, with a spermicide, before sex | You put a latex or silicone device in your vagina, with a spermicide, before sex |
| **How often?** | Every time you have sex | Every time you have sex | Every time you have sex | Every time you have sex | Every time you have sex | Every time you have sex |
| **How does it work?** | Prevents sperm from reaching an egg | Prevents sperm from reaching an egg | Prevents sperm from reaching an egg | Prevents sperm from reaching an egg | Prevents sperm from reaching an egg | Prevents sperm from reaching an egg |
| **When does it start working?** | Immediately | Immediately | Depends on the brand | Immediately | Immediately | Immediately |
| **How many people become pregnant in the first year?** | | | | | | |
| Not always following the instructions: | 18 in 100 people ●●○○○○○○○○ | 21 in 100 people ●●●○○○○○○○ | 28 in 100 people ●●●○○○○○○○ | 12 in 100 people (if never given birth) ●●○○○○○○○○<br>24 in 100 people (if given birth) ●●●○○○○○○○ | 14 in 100 people (if never given birth vaginally) ●●○○○○○○○○<br>29 in 100 people (if given birth vaginally) ●●●○○○○○○○ | 12 in 100 people ●●○○○○○○○○ |
| Always following the instructions: | 2 in 100 people ●○○○○○○○○○ | 5 in 100 people ●○○○○○○○○○ | 18 in 100 people ●●○○○○○○○○ | 9 in 100 people (if never given birth) ●○○○○○○○○○<br>20 in 100 people (if given birth) ●●●○○○○○○○ | *No information available* | 6 in 100 people ●○○○○○○○○○ |
| **What are some of the side effects?** | | | | | | |
| Allergic reaction? | Possible | Possible | Possible | Possible | Possible | Possible |
| Vaginal symptoms (e.g., odor, irritation, infection, vaginitis)? | | | Possible | Possible | Possible | Possible |
| Toxic shock syndrome? | | | | Possible | Possible | Possible |
| **Does it protect against sexually transmitted infections (STIs)?** | Provides the best protection | Provides some protection | No | No | No | No |

For more information, including authors, information sources, and terms of use, see www.rightforme.org/decision

© 2016 Trustees of Dartmouth College | Version 1.0

### Natural Birth Control Methods

right for me

This decision aid is to help you and your health care provider talk about methods of birth control and choose what's right for you. Most people can safely use these methods. Your health care provider can tell you whether these methods are safe for you.

| | | FERTILITY AWARENESS METHODS: | | | |
|---|---|---|---|---|---|
| | **WITHDRAWAL METHOD** | **STANDARD DAYS METHOD®** | **TWODAY METHOD®** | **OVULATION METHOD** | **SYMPTOTHERMAL METHOD** |
| **How is it used?** | During sex, the penis is removed from your vagina and away from your genitals before ejaculation | You monitor the days of your menstrual cycle. You avoid sex on days 8 through 19 of your cycle. | You monitor your cervical secretions. You avoid sex on days you notice secretions and for one day after. | You monitor your cervical secretions. You use a set of rules to know what days to avoid sex. | You monitor your cervical secretions and your body temperature. You use a set of rules to know what days to avoid sex. |
| **How often?** | Every time you have sex | Every day | Every day (checking your secretions two or more times a day) | Every day | Every day |
| **How does it work?** | Prevents sperm from reaching an egg | Prevents sperm from reaching an egg when there is a higher chance of pregnancy | Prevents sperm from reaching an egg when there is a higher chance of pregnancy | Prevents sperm from reaching an egg when there is a higher chance of pregnancy | Prevents sperm from reaching an egg when there is a higher chance of pregnancy |
| **When does it start working?** | Immediately | You can start monitoring the days of your menstrual cycle anytime. It may take time before it is a 'safe day' to have sex. | You can start monitoring your cervical secretions anytime. It may take time before it is a 'safe day' to have sex. | You can start monitoring your cervical secretions anytime. It may take time before it is a 'safe day' to have sex. | You can start monitoring your cervical secretions and your body temperature anytime. It may take time before it is a 'safe day' to have sex. |
| **How many people become pregnant in the first year?** | | | | | |
| Not always following the instructions: | 22 in 100 people ●●●○○○○○○○ | 24 in 100 people ●●●○○○○○○○ | 24 in 100 people ●●●○○○○○○○ | 24 in 100 people ●●●○○○○○○○ | 24 in 100 people ●●●○○○○○○○ |
| Always following the instructions: | 4 in 100 people ●○○○○○○○○○ | 5 in 100 people ●○○○○○○○○○ | 4 in 100 people ●○○○○○○○○○ | 3 in 100 people ●○○○○○○○○○ | Fewer than 1 in 100 people ○○○○○○○○○○ |
| **Are there side effects?** | No | No | No | No | No |
| **Does it protect against sexually transmitted infections (STIs)?** | No | No | No | No | No |

*Note*. The Lactational Amenorrhea Method is another natural birth control method that may be used by some people who are breastfeeding. Your health care provider can tell you about this method.



© 2016 Trustees of Dartmouth College | Version 1.0

### Permanent Birth Control Methods

right for me

This decision aid is to help you and your health care provider talk about methods of birth control and choose what's right for you. Most people can safely use these methods. Your health care provider can tell you whether these methods are safe for you.

| | FEMALE STERILIZATION: | | | MALE STERILIZATION: |
|---|---|---|---|---|
| | BY LAPAROSCOPY | BY MINILAPAROTOMY | BY HYSTEROSCOPY | VASECTOMY |
| **How is it used?** | A health care provider uses instruments inserted through one or two small incisions in your abdomen to be able to see and close your fallopian tubes | A health care provider makes an incision in your abdomen and moves your fallopian tubes up so they can be seen. The provider then closes your tubes. | A health care provider inserts an instrument through your vagina and places small devices in your fallopian tubes. Scar tissue forms around the devices and closes your tubes. | A health care provider makes one or two incisions or a small puncture in the skin of the scrotum and closes the tubes that carry sperm |
| **How often?** | Once | Once | Once | Once |
| **How does it work?** | Prevents sperm from reaching an egg | Prevents sperm from reaching an egg | Prevents sperm from reaching an egg | Prevents sperm from being released |
| **When does it start working?** | Immediately | Immediately | After 3 months, when an X-ray shows that your fallopian tubes are closed | After 2 to 4 months, when a test shows that there are no longer sperm in the semen |
| **How many people become pregnant in the first year?** | | | | |
| Not always following the instructions: | Fewer than 1 in 100 people<br>○○○○○○○○○○ | Fewer than 1 in 100 people<br>○○○○○○○○○○ | Fewer than 1 in 100 people<br>○○○○○○○○○○ | Fewer than 1 in 100 people<br>○○○○○○○○○○ |
| Always following the instructions: | Fewer than 1 in 100 people<br>○○○○○○○○○○ | Fewer than 1 in 100 people<br>○○○○○○○○○○ | Fewer than 1 in 100 people<br>○○○○○○○○○○ | Fewer than 1 in 100 people<br>○○○○○○○○○○ |
| **What are some of the side effects?** | | | | |
| Abdominal cramps or other pain following the procedure? | Possible | Possible | Possible | Possible |
| Dizziness, nausea, vomiting, bleeding, or other symptoms following the procedure? | Possible | Possible | Possible | Possible |
| Minor complication (e.g., infection)? | Possible | Possible | Possible | Possible |
| Major complication (e.g., injury requiring surgery)? | Possible | Possible | Possible | Posible |
| Pain that continues for some time? | Possible | Possible | Possible | Possible |
| **Does it protect against sexually transmitted infections (STIs)?** | No | No | No | No |



© 2016 Trustees of Dartmouth College | Version 1.0

### Emergency Birth Control Methods

right for me

This decision aid is to help you and your health care provider talk about methods of birth control and choose what's right for you.
Most people can safely use these methods. Your health care provider can tell you whether these methods are safe for you.

| | COPPER IUD | ULIPRISTAL PILL | PROGESTIN EMERGENCY PILL | COMBINED PILL |
|---|---|---|---|---|
| **How is it used?** | A health care provider puts a small, T-shaped plastic and copper device in your uterus. You can then use this as regular birth control for up to 10 years. | You swallow one pill | You swallow one pill or you swallow two pills, 12 hours apart (depends on the brand) | You swallow a specific number of combined birth control pills and then repeat 12 hours later. The number of pills needed depends on the brand. |
| **When?** | Within 5 days after unprotected sex | As soon as possible within 5 days after unprotected sex | As soon as possible within 5 days after unprotected sex | As soon as possible within 5 days after unprotected sex |
| **How does it work?** | Releases copper to prevent a pregnancy from occurring | Releases ulipristal acetate to prevent a pregnancy from occurring | Releases progestin to prevent a pregnancy from occurring | Releases progestin and estrogen to prevent a pregnancy from occurring |
| **How effective is it at preventing pregnancy?** | Highly effective – the most effective method | The most effective method after the Copper IUD | As effective as the Ulipristal Pill if taken within 3 days after unprotected sex<br><br>Less effective than the Ulipristal Pill if taken 3 to 5 days after unprotected sex | The least effective method |
| **What are some of the side effects?** | | | | |
| Temporary bleeding changes (e.g., irregular bleeding or spotting, next period not at the expected time)? | | Possible | Possible | Possible |
| Other temporary side effects (e.g., headaches, nausea or vomiting, breast tenderness, abdominal pain, dizziness, fatigue)? | | Possible | Possible | Possible |
| Non-temporary bleeding changes or other side effects? | Possible (More information on the 'Long-Acting Reversible Birth Control Methods' decision aid) | | | |
| **Does it protect against sexually transmitted infections (STIs)?** | No | No | No | No |



© 2016 Trustees of Dartmouth College | Version 1.0

**APPENDIX 5** Right For Me written guidance

right for me

### Frequently Asked Questions

### What is shared decision-making?

The process of health professionals and patients making health decisions together.

© 2016 Trustees of Dartmouth College

### What are the Right For Me Decision Aids?

A set of seven one-page tools that compare birth control methods. They are intended to be used by health professionals during the health care visit. The Decision Aids are available in English and Spanish.

© 2016 Trustees of Dartmouth College

### Who developed the Decision Aids?

Researchers at Dartmouth College and their project partners, including patients, health professionals, and other stakeholders. Decision Aid authors are listed in a Supporting Document, along with a conflict of interest disclosure statement. There is a link to the Supporting Document on each Decision Aid.

© 2016 Trustees of Dartmouth College

### Where did the information come from?

Reviews of scientific evidence, relevant national and international guidelines, and existing patient resources. Information sources are listed in a Supporting Document. There is a link to the Supporting Document on each Decision Aid.

© 2016 Trustees of Dartmouth College

### Where did the questions and format come from?

A survey of over 600 women and health professionals[4], focus groups with patients in three states, and best practice guidelines for decision aid development.

© 2016 Trustees of Dartmouth College

### Who can use the Decision Aids?

Any health professional who provides information or counseling about contraception to patients receiving health care in your clinic.

© 2016 Trustees of Dartmouth College

### With which patients can the Decision Aids be used?

Any English- or Spanish-speaking patients receiving health care in your clinic. You may wish to use the Decision Aids with patients who are considering contraception for the first time, already using a contraceptive method, or interested in changing methods. The reading level of each Decision Aid is 8th grade or less.

© 2016 Trustees of Dartmouth College

### Are the Decision Aids intended to be used by the patient alone?

No. The decision aids are a framework for a conversation with a health professional. Patients may need medical terminology explained. You may also wish to share further information, such as non-contraceptive benefits, other side effects, usage instructions, mechanism of actions, brand names, or off-label use.

© 2016 Trustees of Dartmouth College

### How can I use the Decision Aids with patients?

A demonstration of how to use the Decision Aids is provided in the Decision Aid Training Video. As explained in the Training Video, following three steps may help:

① Explain It ② Give It ③ Use It

© 2016 Trustees of Dartmouth College

### How do I know which Decision Aids to use?

You are encouraged to use as many or as few Decision Aids as makes sense for the patient. The 'Types of Birth Control Methods' Decision Aid may be helpful to use with patients without a lot of background knowledge before narrowing down to a more specific Decision Aid.

© 2016 Trustees of Dartmouth College

### What if some contraceptive methods are unsafe for a patient?

The U.S. Medical Eligibility Criteria for Contraceptive Use and published updates[1–3] provide guidance on who can safely use each method of contraception and may be used to facilitate the provision of individualized information on method safety.

© 2016 Trustees of Dartmouth College

### Can I write on the Decision Aids?

Of course. You are welcome to add or change content to suit the individual patient.

© 2016 Trustees of Dartmouth College

### What does ‘not always following the instructions’ mean?

This is the terminology adopted to convey ‘typical-use effectiveness’ after consultation with patients.

© 2016 Trustees of Dartmouth College

### What does ‘always following the instructions’ mean?

This is the terminology we adopted to convey ‘perfect-use effectiveness’ after consultation with patients.

© 2016 Trustees of Dartmouth College

### What do the dots mean?

To enhance comprehension, effectiveness data are displayed both using raw numbers ("X in 100 people") and using a visual aid based on the following:

| | |
|---|---|
| Less than 1%: | ○○○○○○○○○○ |
| 1% to 9%: | ●○○○○○○○○○ |
| 10% to 19%: | ●●○○○○○○○○ |
| 20% to 29%: | ●●●○○○○○○○ |

© 2016 Trustees of Dartmouth College

### Why are some side effects listed and not others?

Side effects routinely cited in the evidence, guidelines, and existing patient resources reviewed were prioritized for inclusion in the Decision Aids. Due to space constraints, the list of side effects is not exhaustive.

© 2016 Trustees of Dartmouth College

### Under side effects, what does a blank cell mean?

This signifies that the side effect was not attributed to the contraceptive method in the evidence, guidelines, and existing patient resources reviewed.

© 2016 Trustees of Dartmouth College

© 2016 Trustees of Dartmouth College

**APPENDIX 6** Implementation slide deck: Video + Prompt Card

right for me

### Video + Prompt Card

### Introduction

This slide deck is intended to support people in your clinic (administrative staff, health professionals, and others) to implement the Right For Me Video + Prompt Card.

© 2016 Trustees of Dartmouth College

### About

The Right For Me Video is a three-minute video intended to be viewed by patients immediately before the health care visit. It is viewed on an iPad with headphones. The Video is available in English and Spanish, with and without on-screen captions.

The Right For Me Prompt Card is a small card intended to be taken by patients immediately before the health care visit. The Prompt Card is available in English and Spanish.

© 2016 Trustees of Dartmouth College

### Objective

The aim of the Video is to enhance patients' motivation, skills, and self-efficacy to ask health professionals three specific questions:

(1) What are my options?

(2) What are the pros and cons of those options?

(3) How likely are those pros and cons to happen to me?

The aim of the Prompt Card is to remind patients of the three questions presented in the video.

© 2016 Trustees of Dartmouth College

### Audience

The target audience for the Video + Prompt Card is any patient waiting to see a health professional in your clinic (for example, while in the waiting room or after the rooming-in process).

© 2016 Trustees of Dartmouth College

### Development

The Video + Prompt Card were developed by researchers at Dartmouth College and their project partners, based on previous research and extensive consultation with patients, health professionals, and other stakeholders.

© 2016 Trustees of Dartmouth College

### Preview

You can preview the Video + Prompt Card by:

1. Visiting www.rightforme.org
2. Navigating to 'For Clinics' and then 'Video + Prompt Card'
3. Entering the clinic username and password

Your Project Contact will give you the clinic username and password. Please do not share this information with others outside your clinic.

© 2016 Trustees of Dartmouth College

### Implementation

Implementing the Video + Prompt Card is a team effort. The tasks involved are explained below and on the remaining slides.

| What? | Who? |
| --- | --- |
| 1. Supplying the materials | The Right For Me project team |
| 2. Facilitating patients viewing the Video | Administrative staff, health professionals, and/or others |
| 3. Facilitating patients taking the Prompt Card | Administrative staff, health professionals, and/or others |
| 4. Other support tasks | Administrative staff, health professionals, and/or others |

© 2016 Trustees of Dartmouth College

### 1. Supplying the materials

The Right For Me project team

© 2016 Trustees of Dartmouth College

### Right For Me Video

We have supplied your clinic with:

- 2 iPads programmed to view the Video
- 2 sets of headphones
- Cleaning wipes

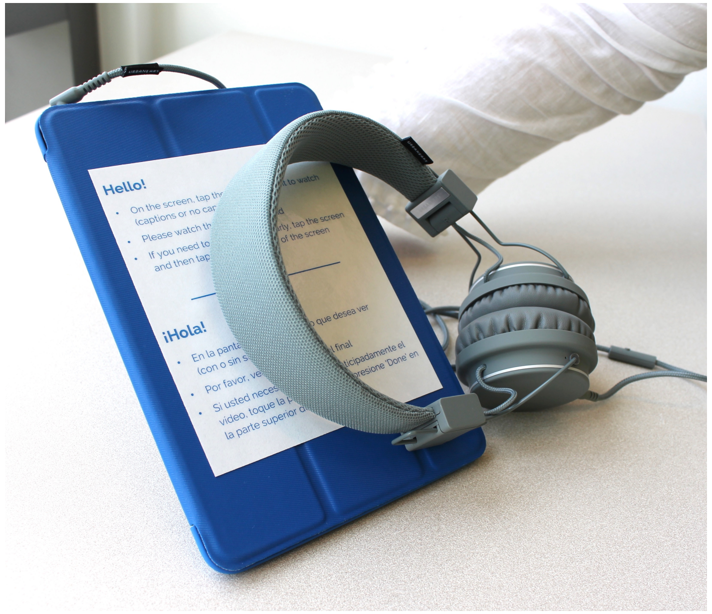


© 2016 Trustees of Dartmouth College

### Right For Me Prompt Card

We have supplied your clinic with:

- 3 display stands of English Prompt Cards
- 1 display stand of Spanish Prompt Cards
- Extra refills of Prompt Cards

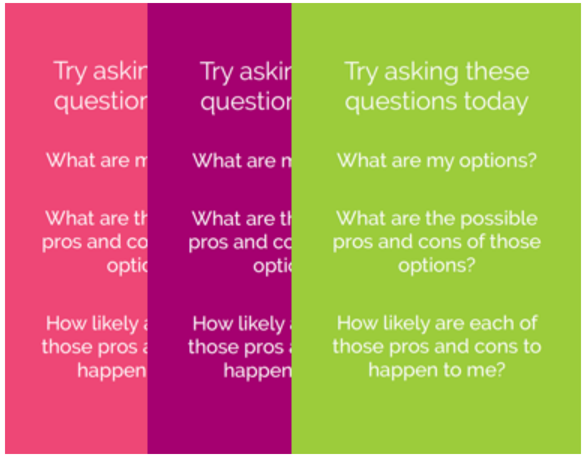


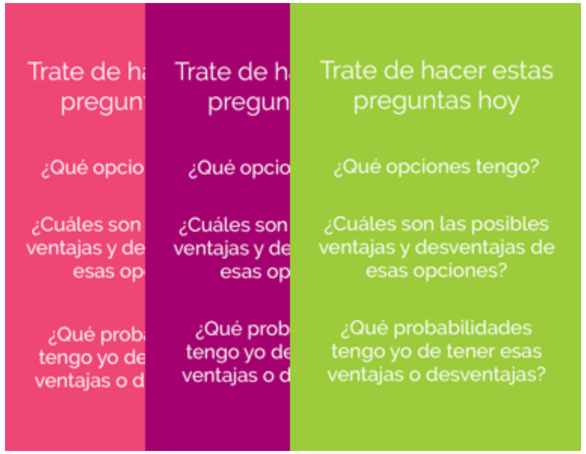


© 2016 Trustees of Dartmouth College

### 2. Facilitating patients viewing the Video

For administrative staff, health professionals, and/or others

© 2016 Trustees of Dartmouth College

### Facilitating Patients Viewing the Video

We encourage you to facilitate patients viewing the Video in a way that works for your clinic. Things you may wish to think about include:

- Who will hand the iPads and headphones to patients?
- When and where will you hand the iPads and headphones to patients?
- What will you say to patients about the Video?
- Who will collect the iPads and headphones from patients?

The next slides provide some practice guidance that may be helpful.

© 2016 Trustees of Dartmouth College

### Guidance for Clinic Staff

Before handing an iPad to a patient, open the cover. You should see four videos displayed on the screen.

If you see that a video is playing, ① tap anywhere on the screen and ② tap 'Done' in the top left corner. Close the cover.

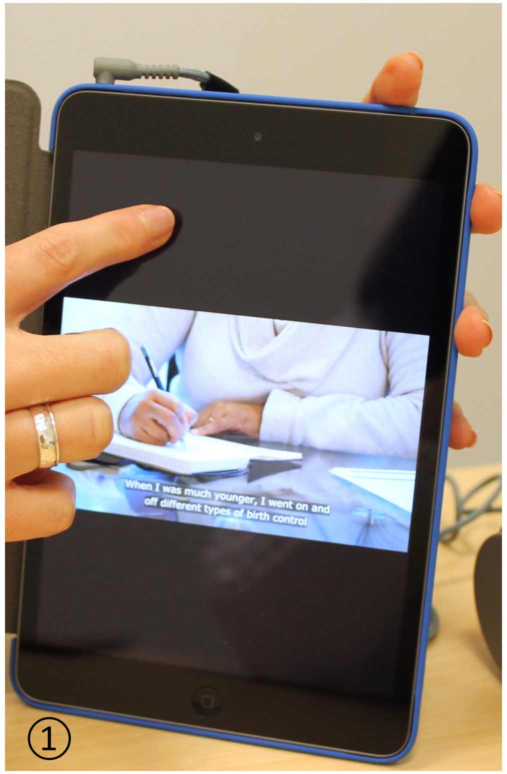


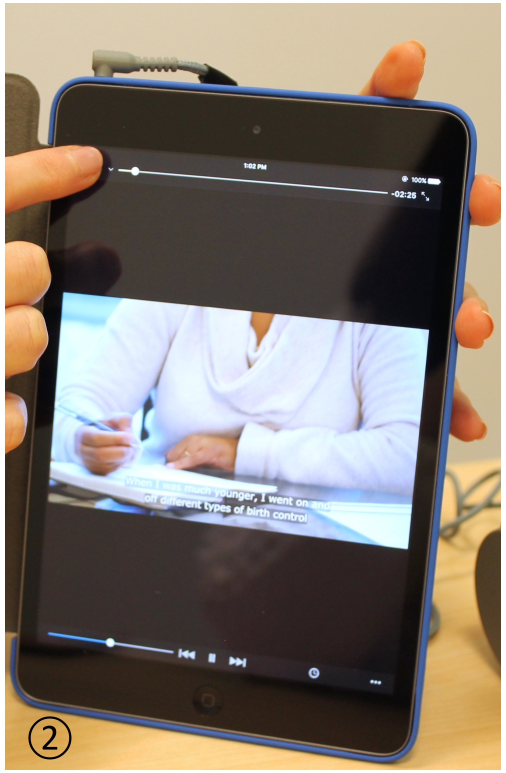


© 2016 Trustees of Dartmouth College

### Guidance for Patients

Basic instructions for patients are provided on the iPad cover in English and Spanish.

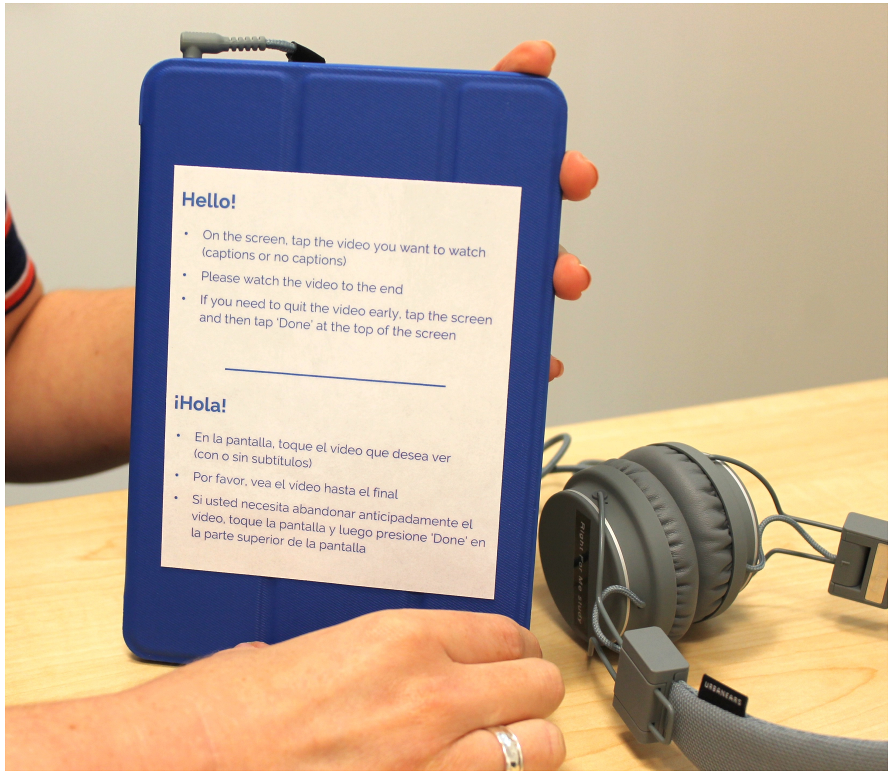


© 2016 Trustees of Dartmouth College

### Guidance for Patients

Patients may choose to watch the Video in English or Spanish, with or without on-screen captions.

Patients should put on the headphones and ① tap the video they want to watch. It will play automatically.

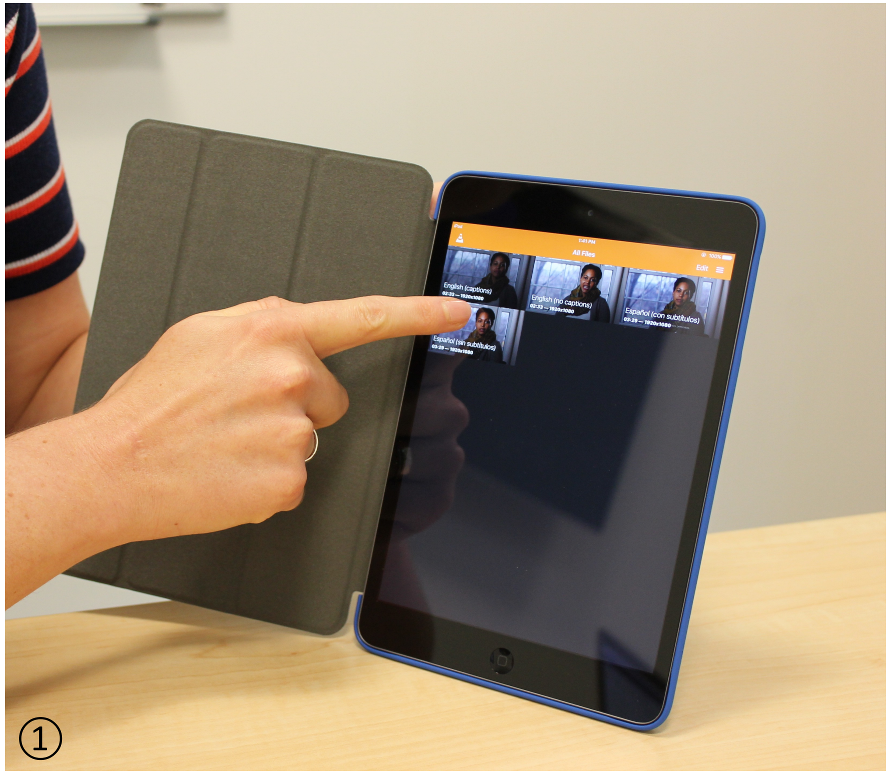


© 2016 Trustees of Dartmouth College

### Guidance for Patients

If a patient wishes to exit a video before it has finished playing, they should ① tap anywhere on the screen and ② tap ‘Done’ in the top left corner.

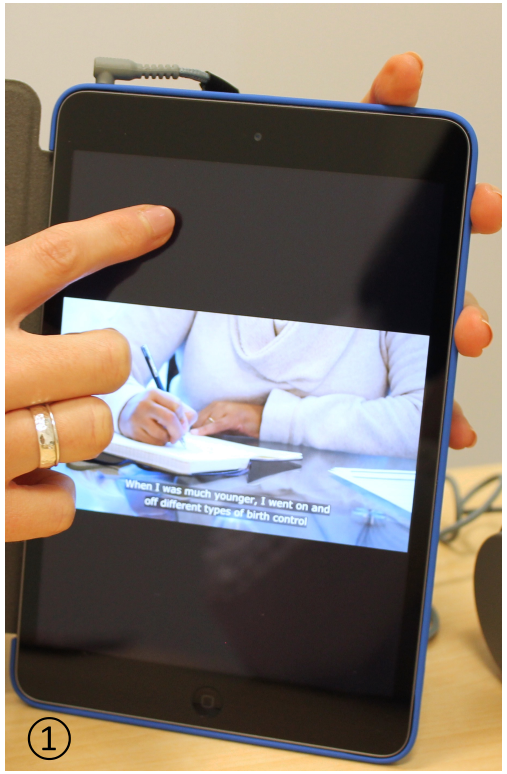


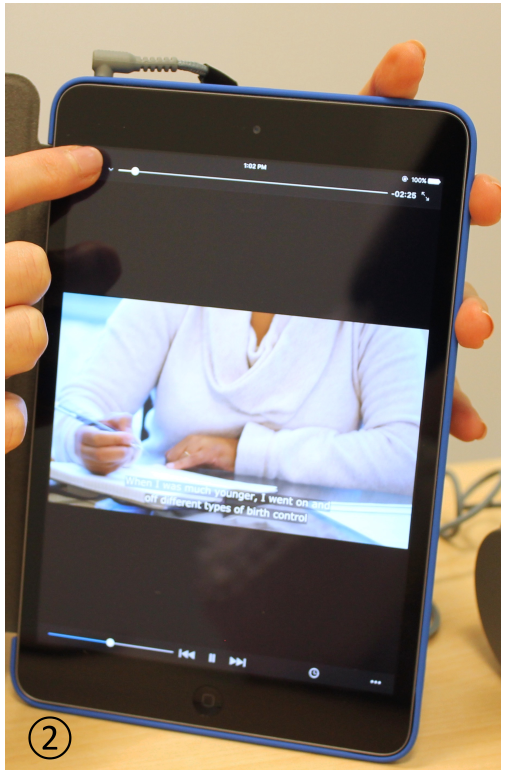


© 2016 Trustees of Dartmouth College

### Guidance for Patients

If a patient wishes to increase or decrease the volume of a video, they should ① press the long up or down button on the right side of the iPad.

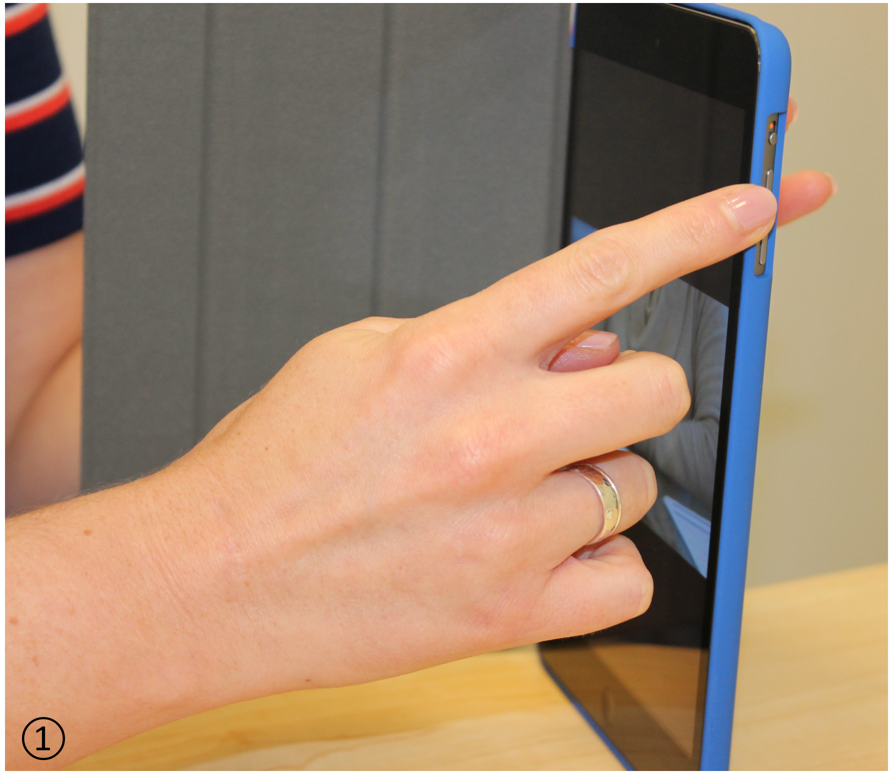


© 2016 Trustees of Dartmouth College

### 3. Facilitating patients taking the Prompt Card

For administrative staff, health professionals, and/or others

© 2016 Trustees of Dartmouth College

### Facilitating Patients Taking the Prompt Card

We encourage you to facilitate patients taking the Prompt Card in a way that works for your clinic. Things you may wish to think about include:

- Will you hand the patient a Prompt Card when you give them the iPad?
- Will you put the Prompt Cards out on display and refer patients to them?
- What will you say to patients about the Prompt Card?

Patients may take the Prompt Card home with them to keep.

© 2016 Trustees of Dartmouth College

### 4. Other support tasks

For administrative staff, health professionals, and/or others

© 2016 Trustees of Dartmouth College

### iPad Management

Please ensure the iPads and headphones are stored securely in the clinic when not in use.

Please charge the iPads as often as needed, including every night. Plug the iPad charger into a wall outlet (not a computer).

© 2016 Trustees of Dartmouth College

### iPad Management

Please check periodically that the iPads remain locked into the Video application. To do this, press the iPad home button at the bottom of the iPad. When you press this, the screen should not change. If the screen changes, please tell your Project Contact.

© 2016 Trustees of Dartmouth College

### Prompt Card Management

Please check periodically to make sure you have enough Prompt Cards. If you are running low on Prompt Cards in English and/or Spanish, please tell your Project Contact. They will ask us to send extra Prompt Cards to your clinic.

© 2016 Trustees of Dartmouth College

Thank you.
Good luck!

© 2016 Trustees of Dartmouth College

**APPENDIX 7** Implementation slide deck: Decision Aids + Training

right for me

### Decision Aids + Training

### Introduction

This slide deck is intended to support people in your clinic (administrative staff, health professionals, and others) to implement the Right For Me Decision Aids + Training.

© 2016 Trustees of Dartmouth College

### About

The Right For Me Decision Aids are a set of seven one-page decision aids on birth control methods. They are intended to be used by health professionals during the health care visit. The Decision Aids are available in English and Spanish.

The Right For Me Decision Aid Training consists of a four-minute Video and Frequently Asked Questions intended to be reviewed by health professionals before beginning to use the Decision Aids.

© 2016 Trustees of Dartmouth College

### Objective

The aim of the Decision Aids is to support health professionals to facilitate shared decision-making about birth control methods with patients in the health care visit.

The aim of the Training Video and Frequently Asked Questions is to enhance health professionals' motivation, skills, and self-efficacy to use the Decision Aids.

© 2016 Trustees of Dartmouth College

### Audience

The target audience for the Decision Aids + Training is any health professional who provides information or counseling about contraception to patients receiving health care in your clinic.

© 2016 Trustees of Dartmouth College

### Development

The Decision Aids + Training were developed by researchers at Dartmouth College and their project partners, based on previous research and extensive consultation with patients, health professionals, and other stakeholders.

© 2016 Trustees of Dartmouth College

### Implementation

Implementing the Decision Aids + Training is a team effort. The tasks involved are explained below and on the remaining slides.

| What? | Who? |
| --- | --- |
| 1. Supplying the materials | The Right For Me project team |
| 2. Taking the Training | Health professionals |
| 3. Using the Decision Aids | Health professionals |
| 4. Other support tasks | Administrative staff, health professionals, and/or others |

© 2016 Trustees of Dartmouth College

### 1. Supplying the materials

The Right For Me project team

© 2016 Trustees of Dartmouth College

### Decision Aids

We have supplied your clinic with:

- Tear-pads of the seven different Decisions Aids in English and Spanish (each set is bound with a rubber band)
- 1 desktop or wall-mounted display stand for each exam room

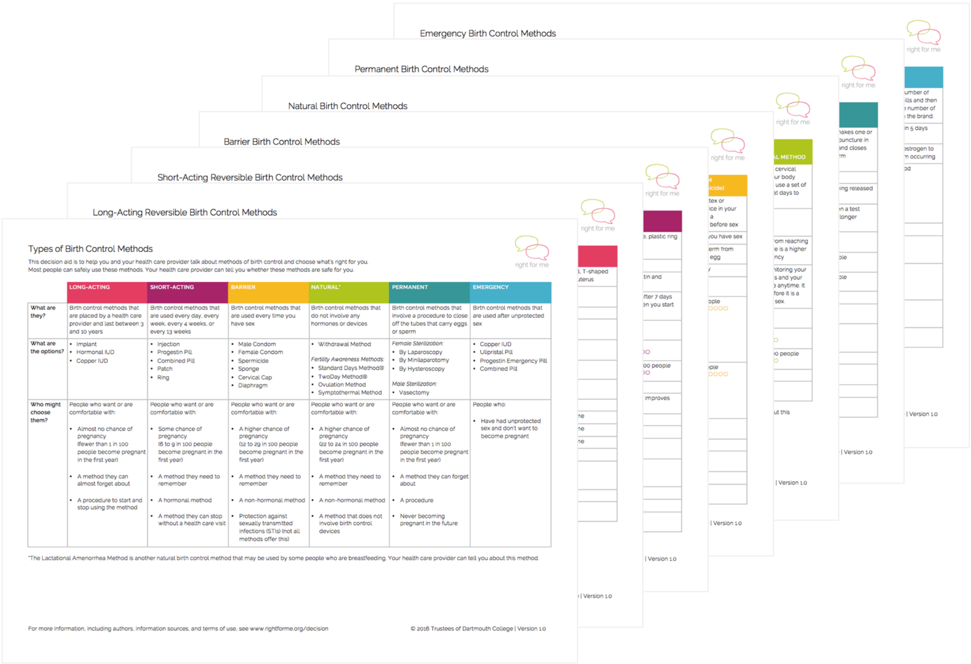


© 2016 Trustees of Dartmouth College

### Training

We have supplied your clinic with online access to:

- The Training Video
- The Frequently Asked Questions

© 2016 Trustees of Dartmouth College

### 2. Taking the Training

For health professionals

© 2016 Trustees of Dartmouth College

### Taking the Training

You can take the Training as often as you wish by:

1. Visiting www.rightforme.org
2. Navigating to 'For Clinics' and then 'Decision Aids + Training'
3. Entering the clinic username and password

Your Project Contact will give you the clinic username and password. Please do not share this information with others outside your clinic.

© 2016 Trustees of Dartmouth College

### 3. Using the Decision Aids

For health professionals

© 2016 Trustees of Dartmouth College

### Using the Decision Aids

All guidance on using the Decision Aids is provided in the Training Video and Frequently Asked Questions.

© 2016 Trustees of Dartmouth College

### 4. Other support tasks

For administrative staff, health professionals, and/or others

© 2016 Trustees of Dartmouth College

### Decision Aid Management

Please place tear-pads of the seven different Decision Aids on display in each exam room.

Please check periodically to make sure that each exam room has enough Decision Aids. If you are running low on Decision Aids in English and/or Spanish, please tell your Project Contact. They will ask us to send extra Decision Aids to your clinic.

© 2016 Trustees of Dartmouth College

Thank you.
Good luck!

© 2016 Trustees of Dartmouth College

#### APPENDIX 8 Strategies for maximising enrolment, retention, and data quality

**Table A2.** Strategies for maximising enrolment, retention, and data quality

| Goal | Strategies |
|---|---|
| Maximise enrolment | • A study name, branding, and engaging recruitment materials designed with patient input<br>• No requirement for documented informed consent or parental consent to participate<br>• Option to participate in the study without a commitment to complete all three surveys<br>• Option to participate in the study without providing name and contact details<br>• Face-to-face training in approved recruitment and data collection processes for clinic staff<br>• Regular feedback to clinic staff on the number of participants enrolling in the study<br>• Participant compensation of a $10 gift card for completing the T1 survey |
| Maximise retention | • Multiple completion modes for the T2 and T3 surveys for participants aged 20 years and older<br>• The use of advance prompts and/or reminders for the T2 and T3 surveys (see Figure A1 below)<br>• Provision of an addressed, reply-paid envelope to facilitate the return of paper surveys<br>• Sending and receiving of T2 and T3 surveys from the research institution (rather than clinic)<br>• Participant compensation of a $10 gift card for completing each of the T2 and T3 surveys |
| Maximise data completeness and quality | • Collection of data via surveys rather than interviews given vulnerability of topic to social desirability bias<br>• Development of surveys and measures by experts in patient-reported outcome and experience measurement in partnership with patients and stakeholders<br>• Online mode of survey completion for the T1 survey to ensure no handling of surveys by clinic staff and thus reinforce the confidentiality of responses<br>• Reassurance in the study information sheet that no health care providers from participating clinics would have access to identified participant-level data<br>• Use of programmed or instructional skips to ensure participants were asked only relevant questions, thereby minimising survey fatigue<br>• Pop-up messages that notified participants of missed questions and invited them to respond to them prior to proceeding to the next page in online surveys<br>• Reminders of clinic name on the tablet computer used to administer the T1 survey<br>• Use of intervention images in items assessing intervention exposure<br>• Participant-specific reminders of the date and clinic of the health care visit and the (previously reported) intended contraceptive method(s) in follow-up surveys<br>• Duplicate data entry for a random subsample of paper surveys to confirm accuracy |

**Figure A1.** Survey invitation and reminder schedule customised to survey mode

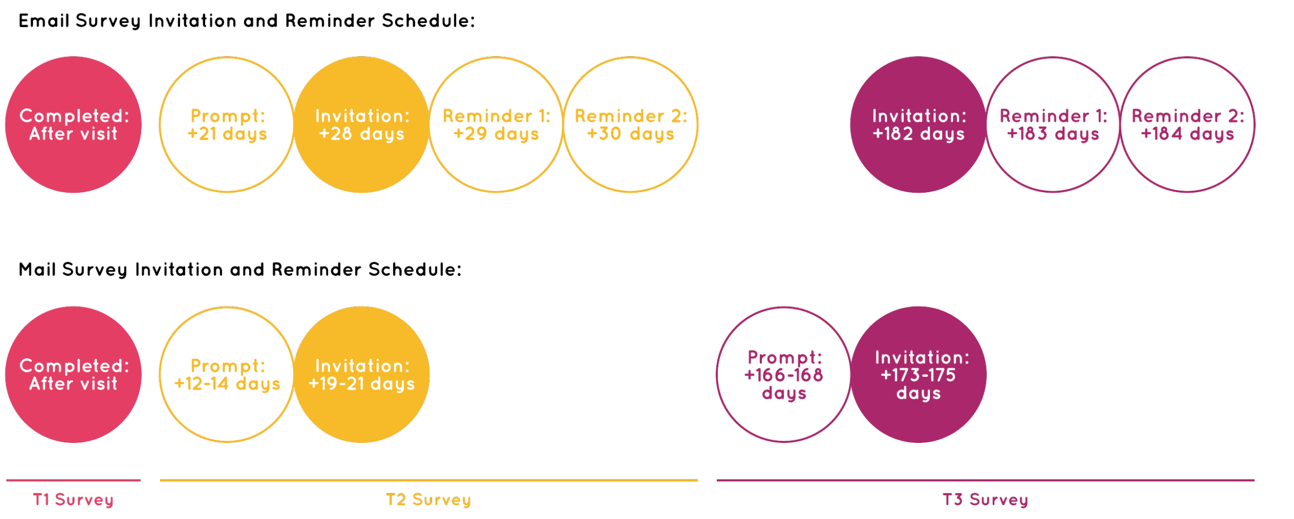

#### APPENDIX 9 Outcomes and measures

**Table A3.** Outcomes and measures

| Outcome | Variable | Coding | Eligible | Excluded |
|---|---|---|---|---|
| **Primary outcome** | | | | |
| Shared decision-making about contraceptive methods | Binary | • Optimal (12 out of 12)<br>• Suboptimal (0-11 out of 12) | Participants who experienced a conversation about contraception in the health care visit | Of the **4,209** participants who were eligible for the study and experienced a conversation about contraception in the health care visit, **21** were excluded due to missing data on shared decision-making about contraceptive methods and a further **108** due to missing data on one or more covariates |
| | Categorical (post-hoc) | • Optimal (12 out of 12)<br>• Moderate (10-11 out of 12)<br>• Low (0-9 out of 12) | | |
| | Continuous (post-hoc) | Range: 0-12 | | |
| **Secondary outcomes** | | | | |
| Conversation about contraception | Binary | • Conversation<br>• No conversation | All participants | Of the **5,038** participants who were eligible for the study, **20** were excluded due to missing data on conversation about contraception and a further **153** due to missing data on one or more covariates |
| | | | ***Prespecified subgroup****:* All participants excluding those not at risk of unintended pregnancy at the time of the health care visit[1] | Of the **5,038** participants who were eligible for the study, **20** were excluded due to missing data on conversation about contraception, **257** due to not meeting subgroup criteria, and a further **142** due to missing data on one or more covariates |
| | | | ***Prespecified subgroup****:* All participants excluding those not at risk of unintended pregnancy at the time of the health care visit[1] and those who reported that they did not want or need to talk about contraception | Of the **5,038** participants who were eligible for the study, **20** were excluded due to missing data on conversation about contraception, **687** due to not meeting subgroup criteria, and a further **133** due to missing data on one or more covariates |
| Satisfaction with conversation about contraception | Binary | • Very satisfied<br>• Somewhat satisfied, neither satisfied nor dissatisfied, somewhat dissatisfied, very dissatisfied | Participants who experienced a conversation about contraception in the index health care visit | Of the **4,209** participants who were eligible for the study and experienced a conversation about contraception in the health care visit, **15** were excluded due to missing data on satisfaction with conversation about contraception and a further **109** due to missing data on one or more covariates |
| Intended contraceptive method(s) | Binary | • ≥1 listed method(s)<br>• 0 listed methods | Participants who experienced a conversation about contraception in the index health care visit | Of the **4,209** participants who were eligible for the study and experienced a conversation about contraception in the health care visit, **26** were excluded due to missing data on intended contraceptive method(s) and a further **101** due to missing data on one or more covariates |
| | | | ***Prespecified subgroup***: Participants who experienced a conversation about contraception in the health care visit excluding those not at risk of unintended pregnancy at the time of the health care visit[1] | Of the **4,209** participants who were eligible for the study and experienced a conversation about contraception in the health care visit, **26** were excluded due to missing data on intended contraceptive method(s), **224** due to not meeting subgroup criteria, and a further **94** due to missing data on one or more covariates. |

| | | | | |
|---|---|---|---|---|
| | | | ***Prespecified subgroup***: Participants who experienced a conversation about contraception in the health care visit excluding those not at risk of unintended pregnancy at the time of the health care visit[1] and those who reported that they did not want or need to use a contraceptive method | n/a |
| Intention to use a highly effective contraceptive method | Binary | • ≥1 highly effective method(s)<br>• 0 highly effective methods | Participants who experienced a conversation about contraception in the index health care visit | Of the **4,209** participants who were eligible for the study and experienced a conversation about contraception in the health care visit, **26** were excluded due to missing data on intended contraceptive method(s) and a further **101** due to missing data on one or more covariates |
| | | | ***Prespecified subgroup***: Participants who experienced a conversation about contraception in the health care visit excluding those not at risk of unintended pregnancy at the time of the health care visit[1] | Of the **4,209** participants who were eligible for the study and experienced a conversation about contraception in the health care visit, **26** were excluded due to missing data on intended contraceptive method(s), **224** due to not meeting subgroup criteria, and a further **94** due to missing data on one or more covariates. |
| | | | ***Prespecified subgroup***: Participants who experienced a conversation about contraception in the health care visit excluding those not at risk of unintended pregnancy at the time of the health care visit[1] and those who reported that they did not want or need to use a contraceptive method | Of the **4,209** participants who were eligible for the study and experienced a conversation about contraception in the health care visit, **26** were excluded due to missing data on intended contraceptive method(s), **317** due to not meeting subgroup criteria, and a further **87** due to missing data on one or more covariates. |
| Values concordance of intended contraceptive method(s) | Binary | • Very confident<br>• Confident, somewhat confident, not very confident, not at all confident | Participants who experienced a conversation about contraception in the health care visit and intended to use ≥1 contraceptive method(s) | **T1**: Of the **3,772** participants who were eligible for the study, experienced a conversation about contraception in the health care visit, and intended to use ≥1 contraceptive method(s), **5** were excluded due to missing data on values concordance of intended contraceptive method(s) and a further **79** due to missing data on one or more covariates<br><br>**T2,T3**: Of the **3,772** participants who were eligible for the study, experienced a conversation about contraception in the health care visit, and intended to use ≥1 contraceptive method(s), **1,301** were excluded due to not giving permission to be contacted for follow-up data collection, **1,213** due to not participating at follow-up, **1** due to missing data on values concordance of intended contraceptive method(s), and a further **5** due to missing data on one or more covariates |
| Decision regret about intended contraceptive method(s) | Continuous | Range: 0-4 | Participants who experienced a conversation about contraception in the index health care visit and intended to use ≥1contraceptive method(s) | Of the **3,772** participants who were eligible for the study, experienced a conversation about contraception in the health care visit, and intended to use ≥1 contraceptive method(s), **1,301** were excluded due to not giving permission to be contacted for follow-up data collection, **1,213** due to not participating at follow-up, **3** due to missing data on decision regret about intended contraceptive method(s), and a further **5** due to missing data on one or more covariates |

| | | | | |
|---|---|---|---|---|
| Contraceptive method(s) used | Binary | • ≥1 listed method(s)<br>• 0 listed methods | Participants who experienced a conversation about contraception in the index health care visit and intended to use ≥1 contraceptive method(s) | n/a |
| Use of a highly effective contraceptive method | Binary | • ≥1 highly effective method(s)<br>• 0 highly effective methods | Participants who experienced a conversation about contraception in the index health care visit and intended to use ≥1contraceptive method(s) | n/a |
| Use of intended contraceptive method(s) | Binary | • Perfect concordance of intended and used method(s)<br>• Imperfect concordance of intended and used method(s) | Participants who experienced a conversation about contraception in the index health care visit and intended to use ≥1contraceptive method(s) | Of the **3,772** participants who were eligible for the study, experienced a conversation about contraception in the health care visit, and intended to use ≥1 contraceptive method(s), **1,301** were excluded due to not giving permission to be contacted for follow-up data collection, **1,213** due to not participating at follow-up, **1** due to missing data on use of intended contraceptive method(s), and a further **5** due to missing data on one or more covariates |
| Adherence to contraceptive method(s) used | Binary | • Perfect adherence to ≥1 method<br>• Perfect adherence to 0 methods | Participants who experienced a conversation about contraception in the index health care visit, intended to use ≥1 contraceptive method(s), and used ≥1 contraceptive method(s) | Of the **1,212** participants who were eligible for the study, experienced a conversation about contraception in the health care visit, intended to use ≥1 contraceptive method(s), and used ≥1 contraceptive method(s), **2** were excluded due to missing data on adherence to contraceptive method(s) used and a further **5** due to missing data on one or more covariates |
| Satisfaction with contraceptive method(s) used | Binary | • Very satisfied<br>• Somewhat satisfied, neither satisfied nor dissatisfied, somewhat dissatisfied, very dissatisfied | Participants who experienced a conversation about contraception in the index health care visit, intended to use ≥1 contraceptive method(s), and used either none of the listed contraceptive methods or ≥1 contraceptive method(s) | Of the **1,257** participants who were eligible for the study, experienced a conversation about contraception in the health care visit, intended to use ≥1 contraceptive method(s), and used either none of the listed contraceptive methods or ≥1 contraceptive method(s), **3** were excluded due to missing data on satisfaction with contraceptive method(s) used and a further **5** due to missing data on one or more covariates |
| Unintended pregnancy (timing preferences) | Binary | • ≥1 unintended pregnancies<br>• 0 unintended pregnancies | Participants who experienced a conversation about contraception in the index health care visit and intended to use ≥1contraceptive method(s) | n/a |
| Unintended pregnancy (pregnancy seeking) | Binary | • ≥1 unintended pregnancies<br>• 0 unintended pregnancies | Participants who experienced a conversation about contraception in the index health care visit and intended to use ≥1contraceptive method(s) | n/a |
| Unwelcome pregnancy | Binary | • ≥1 unwelcome pregnancies<br>• 0 unwelcome pregnancies | Participants who experienced a conversation about contraception in the index health care visit and intended to use ≥1contraceptive method(s) | n/a |
| **Process outcomes** | | | | |
| Intervention exposure[2] | Binary[3] | • Intended exposure<br>• Unintended exposure | Participants with data on at least one study outcome[4] | n/a |
| Acceptability of interventions[2] | Binary | • Would recommend to a friend<br>• Would not recommend to a friend | Participants exposed to the relevant intervention component as intended[5] | n/a |

*Notes*. Grey cells signify planned analyses that could not be conducted. [1]Participants were considered not at risk of unintended pregnancy if they reported they were pregnant, were trying to get pregnant, were born without ovaries or a uterus, had had their ovaries or uterus removed, had entered menopause, were infertile, or did not plan to have vaginal sex with a person who produces sperm. [2]Only descriptive analyses of these outcomes were conducted (see *Analysis*). [3]Exposure outcomes were planned as binary variables. Because only descriptive analyses were conducted, categorical data are also presented. [4]Each outcome was measured among all participants and is reported among participants with data on at least one study outcome. [5]Each outcomes was measured among participants with any exposure to the relevant intervention component and is reported among participants exposed to the relevant intervention component as intended.

#### APPENDIX 10 Impact of patient and public involvement

**Table A4.** Impact of patient and public involvement

| Domain | Illustrative impacts on decisions and materials |
|---|---|
| Objectives | • Decision to include the prompt card in the video intervention strategy<br>• Refinement of conceptualisation of some of the secondary outcomes (e.g., for values concordance of intended contraceptive method(s), to focus on values concordance as perceived by the patient and not by a third party)<br>• Addition of the secondary outcome of unwelcome pregnancy<br>• Planned subgroup analyses for some secondary outcomes |
| Interventions | • Content domains and content of the decision aids<br>• Visual presentation of textual and numerical information in the decision aids<br>• Language used in the decision aids<br>• Content of the script for the (provider) training video<br>• Refinement of the wording of one of the questions advocated by the video and prompt card to “pros and cons”<br>• Creation of a live rather than animated (patient) video that featured an authentic patient story<br>• Content of the script for the (patient) video<br>• Creation of versions of the (patient) video with on-screen captions for patients with hearing impairments |
| Methods | • Creation of a patient-friendly study name (and the specific name, ‘Right For Me’)<br>• Decision not to create eligibility criteria pertaining to current gender identity<br>• Language used to communicate eligibility criteria to prospective participants<br>• Language used in surveys and in new or adapted patient-reported outcome measures (including question stem and response options for the Measure of Alignment of Choices (MATCH))<br>• Conceptualisation and design of strategies for enhancing recruitment, retention, and data quality (including materials to provide regular feedback to clinic staff on the number of participants enrolling in the study)<br>• Inclusion of four-week follow-up data collection in addition to six-month follow-up data collection<br>• Coding of variables for heterogeneity of treatment effects analyses |
| Results | • Interpretation of findings |

#### APPENDIX 11 Protocol refinements and deviations

**Table A5.** Protocol refinements and deviations

| Method | Change or addition |
|---|---|
| Study setting | During the study, we became aware that some clinics used staff members from affiliated clinics that were also participating in the study to temporarily cover staff shortages. Attempts were made by clinics themselves to avoid any contamination arising from this. |
| Participants | Some participants (n=29) were believed to have enrolled in the study more than once, despite passing eligibility screening. Suspected duplicate participation attempts were retrospectively classified as ineligible by researchers.<br>During the study, after publication of the study protocol, we devised eligibility rules pertaining to the required timing of completion of the T2 and T3 surveys relative to the target completion date. These rules required T2 surveys to be completed within one week before or after the target completion date and T3 surveys to be completed within two weeks before or after the target completion date. |
| Study outcomes | In our study protocol, we did not specify precisely how the outcomes of intended contraceptive method(s) or contraceptive method(s) used would be coded. Given the diversity in participant responses to items assessing these two outcomes, we elected to code these outcomes as binary variables. |
| Study time frame | Due to delays, the enrolment of participants and administration of the T1 survey among the pre-implementation cohort began after April 1, 2016 in two clinics, and the implementation of the intervention(s) and administration of the T1 survey among the post-implementation cohort began after July 1, 2016 in one clinic.<br>Enrolment of participants and administration of the T1 survey among the post-implementation cohort continued until December 30, 2016 in one clinic. |
| Data collection | Due to weekends, public holidays, or other planned staff absences, there were some minor deviations from the planned survey mailing timeframe. |

#### APPENDIX 12 Participant flow

**Figure A2.** Flowchart of participants who consented to the study by trial arm

| | **Video** (4 clinics) | **Decision Aids** (4 clinics) | **Dual** (4 clinics) | **Usual Care** (4 clinics) | **All** (16 clinics) |
|---|---|---|---|---|---|
| Consented to participate in study: | **n=1,340**<br>Pre: n=401<br>Post: n=939 | **n=1,218**<br>Pre: n=504<br>Post: n=714 | **n=1,502**<br>Pre: n=469<br>Post: n=1,033 | **n=1,329**<br>Pre: n=420<br>Post: n=909 | **n=5,389**<br>Pre: n=1,794<br>Post: n=3,595 |
| Ineligible for study: | n=84 | n=75 | n=109 | n=83 | n=351 |
| Eligible for study: | **n=1,256**<br>Pre: n=374<br>Post: n=882 | **n=1,143**<br>Pre: n=471<br>Post: n=672 | **n=1,393**<br>Pre: n=445<br>Post: n=948 | **n=1,246**<br>Pre: n=401<br>Post: n=845 | **n=5,038**<br>Pre: n=1,691<br>Post: n=3,347 |
| No conversation about contraception: | n=222 | n=91 | n=239 | n=277 | n=829 |
| Had conversation about contraception: | **n=1,034**<br>Pre: n=306<br>Post: n=728 | **n=1,052**<br>Pre: n=420<br>Post: n=632 | **n=1,154**<br>Pre: n=358<br>Post: n=796 | **n=969**<br>Pre: n=307<br>Post: n=662 | **n=4,209**<br>Pre: n=1,391<br>Post: n=2,818 |
| Not in primary outcome analyses: | n=39 | n=21 | n=37 | n=32 | n=129 |
| In primary outcome analyses: | **n=995**<br>Pre: n=296<br>Post: n=699 | **n=1,031**<br>Pre: n=414<br>Post: n=617 | **n=1,117**<br>Pre: n=345<br>Post: n=772 | **n=937**<br>Pre: n=299<br>Post: n=638 | **n=4,080**<br>Pre: n=1,354<br>Post: n=2,726 |
| Eligible to participate at follow-up: | **n=935**<br>Pre: n=272<br>Post: n=663 | **n=962**<br>Pre: n=382<br>Post: n=580 | **n=1,022**<br>Pre: n=329<br>Post: n=693 | **n=853**<br>Pre: n=270<br>Post: n=583 | **n=3,772**<br>Pre: n=1,253<br>Post: n=2,519 |
| No permission to be contacted: | n=356 | n=336 | n=320 | n=289 | n=1,301 |
| Permission to be contacted: | **n=579**<br>Pre: n=179<br>Post: n=400 | **n=626**<br>Pre: n=249<br>Post: n=377 | **xn=702**<br>Pre: n=204<br>Post: n=498 | **n=564**<br>Pre: n=164<br>Post: n=400 | **n=2,471**<br>Pre: n=796<br>Post: n=1,675 |
| Did not participate at follow-up: | n=276 | n=348 | n=329 | n=260 | n=1,213 |
| Participated at follow-up: | **n=303**<br>Pre: n=100<br>Post: n=203 | **n=278**<br>Pre: n=115<br>Post: n=163 | **n=373**<br>Pre: n=117<br>Post: n=256 | **n=304**<br>Pre: n=102<br>Post: n=202 | **n=1,258**<br>Pre: n=434<br>Post: n=824 |

**Table A6.** Intervention effects on shared decision-making about contraceptive methods (continuous scoring method): differences between pre-implementation and post-implementation cohorts and difference-in-differences analyses

| | **Pre-implementation cohort** M (SD) | **Post-implementation cohort** M (SD) | **Difference between pre-implementation [REF] and post-implementation cohorts** ALSM[1] (95% CI) |
|---|---|---|---|
| **All participants (n=4,080)** | | | |
| Video (n=995) | 10.51 (2.20) | 10.61 (1.96) | -0.02 (-0.26, 0.23) |
| Decision aids (n=1,031) | 10.94 (1.80) | 11.16 (1.51) | 0.14 (-0.09, 0.37) |
| Dual (n=1,117) | 10.89 (1.87) | 10.81 (1.82) | -0.00 (-0.24, 0.23) |
| Usual care (n=937) | 10.98 (1.57) | 10.69 (1.95) | **-0.29 (-0.54, -0.05)** |

| | **Difference-in-differences** ALSM[1] (95% CI) | **P-value unadjusted for multiple comparisons** | **P-value adjusted for multiple comparisons** |
|---|---|---|---|
| **All participants (n=4,080)** | | | |
| Video vs. Usual care [REF] | 0.28 (-0.07, 0.63) | 0.12 | 0.49 |
| Decision aids vs. Usual care [REF] | **0.44 (0.10, 0.77)** | **0.01** | 0.09 |
| Dual vs. Video [REF] | 0.01 (-0.33, 0.35) | 0.95 | 1.00 |
| Dual vs. Decision aids [REF] | -0.15 (-0.47, 0.18) | 0.38 | 0.86 |

*Notes.* ALSM = Adjusted Least Squares Mean, CI = Confidence Interval, M = Mean, REF = Reference group, SD = Standard Deviation.
[1]Analyses adjusted for age, health insurance coverage, health literacy, educational attainment, ethnicity, race, number of pregnancies, number of births, number of abortions, number of miscarriages, previous contraceptive method(s), and survey language.

**Table A7.** Intervention effects on shared decision-making about contraceptive methods (categorical scoring method): differences between pre-implementation and post-implementation cohorts and difference-in-differences analyses

| | **Pre-implementation cohort** | | | **Post-implementation cohort** | | | **Difference between pre-implementation [REF] and post-implementation cohorts** |
|---|---|---|---|---|---|---|---|
| | % low | % moderate | % optimal | % low | % moderate | % optimal | ACOR[1] (95% CI) |
| **All participants (n=4,080)** | | | | | | | |
| Video (n=995) | 31.8 | 12.5 | 55.7 | 30.9 | 13.0 | 56.1 | 0.92 (0.70, 1.22) |
| Decision aids (n=1,031) | 22.7 | 11.6 | 65.7 | 18.5 | 12.3 | 69.2 | 1.11 (0.85, 1.45) |
| Dual (n=1,117) | 23.5 | 12.2 | 64.3 | 25.4 | 15.2 | 59.5 | 0.90 (0.69, 1.18) |
| Usual care (n=937) | 23.7 | 12.7 | 63.5 | 28.1 | 15.5 | 56.4 | **0.75 (0.57, 0.99)** |

| | **Difference-in-differences** ACOR[1] (95% CI) | **P-value unadjusted for multiple comparisons** | **P-value adjusted for multiple comparisons** |
|---|---|---|---|
| **All participants (n=4,080)** | | | |
| Video vs. Usual care [REF] | 1.23 (0.83, 1.83) | 0.30 | 0.78 |
| Decision aids vs. Usual care [REF] | **1.48 (1.01, 2.19)** | **0.05** | 0.26 |
| Dual vs. Video [REF] | 0.98 (0.67, 1.44) | 0.91 | 1.00 |
| Dual vs. Decision aids [REF] | 0.81 (0.56, 1.19) | 0.28 | 0.76 |

*Notes.* ACOR = Adjusted Cumulative Odds Ratio, CI = Confidence Interval, REF = Reference group. [1]Analyses adjusted for age, health insurance coverage, health literacy, educational attainment, ethnicity, race, number of pregnancies, number of births, number of abortions, number of miscarriages, previous contraceptive method(s), and survey language.

**Table A8.** P-values for three-way interactions to assess heterogeneity of treatment effects on shared decision-making about contraceptive methods

| Variable[1] | Variable levels | P-value for three-way interaction between arm, cohort, and variable[2] |
|---|---|---|
| Age | 15 to 24 years, 25 to 49 years | 0.66 |
| Health insurance coverage | None, Public only, Other | 0.06 |
| Health literacy | Limited, Adequate | **<0.0001** |
| Educational attainment | High school graduate or less, Some college or more | 0.58 |
| Ethnicity and race | White alone and not of Hispanic, Latino, or Spanish origin, Of Hispanic, Latino, or Spanish origin and/or another race(s) | 0.61 |
| Number of pregnancies | None, One or more | 0.39 |
| Number of births | None, One or more | 0.22 |
| Number of abortions | None, One or more | 0.77 |
| Number of miscarriages | None, One or more | 0.44 |
| Previous contraceptive method(s) | None, Less effective, Highly effective | 0.85 |

*Notes*. [1]For the variables of age, health insurance coverage, health literacy, educational attainment, ethnicity and race, and previous contraceptive method(s), analyses adjusted for age, health insurance coverage, health literacy, educational attainment, ethnicity, race, number of pregnancies, number of births, number of abortions, number of miscarriages, previous contraceptive method(s), and survey language (except the variable itself). For the variables of number of pregnancies, number of births, number of abortions, and number of miscarriages, analyses adjusted for age, health insurance coverage, health literacy, educational attainment, ethnicity, race, previous contraceptive method(s), and survey language. [2]P-values were compared to a critical value of .005 to account for the consequences of multiple comparisons.

**Table A9.** Intervention effects on secondary outcomes: differences between pre-implementation and post-implementation cohorts

| | **Pre-implementation cohort** % | **Post-implementation cohort** % | **Difference between pre-implementation [REF] and post-implementation cohorts** AOR[1] (95% CI) |
|---|---|---|---|
| CONVERSATION ABOUT CONTRACEPTION (n=4,865) | | | |
| Video (n=1,202) | 82.5 | 83.5 | 0.86 (0.60, 1.24) |
| Decision aids (n=1,116) | 89.4 | 94.5 | 1.58 (0.97, 2.55) |
| Dual (n=1,342) | 81.0 | 84.4 | 1.35 (0.94, 1.93) |
| Usual care (n=1,205) | 76.5 | 78.6 | 1.41 (0.99, 2.03) |
| ***Prespecified subgroup excluding those not at risk of unintended pregnancy (n=4,619)*** | | | |
| Video (n=1,173) | 84.4 | 85.6 | 0.86 (0.58, 1.26) |
| Decision aids (n=1,097) | 91.2 | 96.0 | 1.69 (0.98, 2.92) |
| Dual (n=1,212) | 92.5 | 92.1 | 1.05 (0.64, 1.72) |
| Usual care (n=1,137) | 81.0 | 83.3 | 1.43 (0.97, 2.12) |
| ***Prespecified subgroup excluding those not at risk of unintended pregnancy and those who reported that they did not want or need to talk about contraception (n=4,198)*** | | | |
| Video (n=1,017) | 98.0 | 98.5 | 1.18 (0.42, 3.33) |
| Decision aids (n=1,038) | 98.8 | 99.7 | 3.26 (0.61, 17.35) |
| Dual (n=1,148) | 96.6 | 97.7 | 1.90 (0.86, 4.21) |
| Usual care (n=995) | 92.9 | 95.1 | 1.80 (0.92, 3.51) |
| SATISFACTION WITH CONVERSATION ABOUT CONTRACEPTION (n=4,085) | | | |
| Video (n=1,000) | 85.9 | 89.2 | 1.19 (0.78, 1.83) |
| Decision aids (n=1,031) | 93.5 | 94.8 | 1.21 (0.70, 2.08) |
| Dual (n=1,118) | 90.4 | 89.8 | 1.02 (0.65, 1.59) |
| Usual care (n=936) | 90.9 | 91.4 | 1.09 (0.66, 1.79) |
| INTENDED CONTRACEPTIVE METHOD(S) (n=4,082) | | | |
| Video (n=998) | 90.6 | 91.4 | 1.11 (0.65, 1.89) |
| Decision aids (n=1031) | 91.8 | 91.9 | 0.86 (0.51, 1.46) |
| Dual (n=1117) | 92.4 | 88.0 | 0.67 (0.39, 1.16) |
| Usual care (n=936) | 88.6 | 89.2 | 1.13 (0.67, 1.90) |
| ***Prespecified subgroup excluding those not at risk of unintended pregnancy (n=3,865)*** | | | |
| Video (n=955) | 95.1 | 95.4 | 0.99 (0.48, 2.05) |
| Decision aids (n=980) | 96.7 | 96.6 | 0.73 (0.34, 1.60) |
| Dual (n=1,045) | 97.2 | 94.7 | 0.54 (0.23, 1.24) |
| Usual care (n=885) | 94.0 | 94.2 | 1.55 (0.75, 3.18) |
| INTENTION TO USE A HIGHLY EFFECTIVE CONTRACEPTIVE METHOD (n=4,082) | | | |
| Video (n=998) | 26.2 | 27.3 | 1.01 (0.68, 1.51) |
| Decision aids (n=1,031) | 30.4 | 31.1 | 1.10 (0.77, 1.56) |
| Dual (n=1,117) | 39.5 | 31.4 | 0.74 (0.52, 1.05) |
| Usual care (n=936) | 37.5 | 38.1 | 1.04 (0.71, 1.53) |
| ***Prespecified subgroup excluding those not at risk of unintended pregnancy (n=3,865)*** | | | |
| Video (n=955) | 27.5 | 28.5 | 0.95 (0.63, 1.43) |
| Decision aids (n=980) | 32.1 | 32.7 | 1.14 (0.79, 1.63) |
| Dual (n=1,045) | 41.6 | 33.8 | 0.72 (0.50, 1.04) |
| Usual care (n=885) | 39.7 | 40.3 | 1.07 (0.72, 1.60) |
| ***Prespecified subgroup excluding those not at risk of unintended pregnancy and those who reported that they did not want or need to use a contraceptive method (n=3,779)*** | | | |
| Video (n=938) | 27.9 | 29.0 | 0.99 (0.65, 1.50) |

| | | | |
|---|---|---|---|
| Decision aids (n=961) | 32.6 | 33.4 | 1.17 (0.81, 1.69) |
| Dual (n=1,027) | 42.2 | 34.5 | 0.71 (0.49, 1.04) |
| Usual care (n=853) | 41.3 | 41.8 | 1.05 (0.69, 1.57) |
| VALUES CONCORDANCE OF INTENDED CONTRACEPTIVE METHOD(S) | | | |
| **Immediately after the health care visit (n=3,688)** | | | |
| Video (n=910) | 54.8 | 53.8 | 0.91 (0.67, 1.22) |
| Decision aids (n=947) | 66.8 | 67.0 | 0.98 (0.73, 1.30) |
| Dual (n=998) | 63.8 | 64.9 | 1.05 (0.79, 1.40) |
| Usual care (n=833) | 62.3 | 59.3 | 0.88 (0.64, 1.19) |
| **Four weeks after the health care visit (n=1,252)** | | | |
| Video (n=264) | 43.4 | 44.8 | 0.91 (0.53, 1.59) |
| Decision aids (n=232) | 54.0 | 46.2 | 0.71 (0.41, 1.23) |
| Dual (n=336) | 57.5 | 52.2 | 0.88 (0.55, 1.43) |
| Usual care (n=262) | 44.8 | 49.7 | 1.27 (0.74, 2.17) |
| **Six months after the health care visit (n=1,252)** | | | |
| Video (n=238) | 50.6 | 44.7 | 0.75 (0.42, 1.33) |
| Decision aids (n=215) | 47.1 | 35.9 | 0.58 (0.32, 1.03) |
| Dual (n=275) | 50.0 | 49.7 | 1.06 (0.62, 1.83) |
| Usual care (n=242) | 48.2 | 49.7 | 1.13 (0.65, 1.96) |
| USE OF INTENDED CONTRACEPTIVE METHOD(S) | | | |
| **Four weeks after the health care visit (n=1,252)** | | | |
| Video (n=264) | 57.8 | 64.1 | 1.21 (0.70, 2.10) |
| Decision aids (n=232) | 56.0 | 56.8 | 1.03 (0.60, 1.78) |
| Dual (n=337) | 61.7 | 61.3 | 1.00 (0.62, 1.63) |
| Usual care (n=262) | 58.6 | 63.4 | 1.24 (0.72, 2.12) |
| **Six months after the health care visit (n=1,252)** | | | |
| Video (n=237) | 59.2 | 56.5 | 0.84 (0.47, 1.48) |
| Decision aids (n=215) | 59.8 | 46.9 | 0.59 (0.33, 1.05) |
| Dual (n=275) | 57.5 | 61.0 | 1.22 (0.71, 2.10) |
| Usual care (n=242) | 50.6 | 55.3 | 1.32 (0.76, 2.27) |
| ADHERENCE TO CONTRACEPTIVE METHOD(S) USED | | | |
| **Four weeks after the health care visit (n=1,205)** | | | |
| Video (n=255) | 74.4 | 77.4 | 1.19 (0.61, 2.32) |
| Decision aids (n=219) | 89.2 | 77.0 | **0.43 (0.19, 0.97)** |
| Dual (n=316) | 84.5 | 83.6 | 1.13 (0.57, 2.24) |
| Usual care (n=250) | 83.5 | 84.8 | 1.00 (0.47, 2.13) |
| **Six months after the health care visit (n=1,205)** | | | |
| Video (n=225) | 69.5 | 75.0 | 1.52 (0.77, 2.99) |
| Decision aids (n=198) | 94.0 | 80.9 | **0.29 (0.10, 0.84)** |
| Dual (n=259) | 88.3 | 79.7 | 0.58 (0.26, 1.32) |
| Usual care (n=230) | 87.7 | 84.6 | 0.69 (0.30, 1.60) |
| SATISFACTION WITH CONTRACEPTIVE METHOD(S) USED | | | |
| **Four weeks after the health care visit (n=1,249)** | | | |
| Video (n=264) | 56.6 | 70.2 | 1.58 (0.90, 2.78) |
| Decision aids (n=231) | 62.0 | 62.6 | 1.09 (0.62, 1.90) |
| Dual (n=336) | 67.3 | 69.0 | 1.15 (0.70, 1.91) |
| Usual care (n=262) | 64.4 | 73.1 | 1.55 (0.88, 2.74) |
| **Six months after the health care visit (n=1,249)** | | | |
| Video (n=237) | 65.8 | 112 (69.6) | 1.13 (0.62, 2.06) |
| Decision aids (n=214) | 69.0 | 81 (63.8) | 0.82 (0.45, 1.49) |

| | | | |
|---|---|---|---|
| Dual (n=275) | 88.8 | 138 (70.8) | **0.32 (0.15, 0.72)** |
| Usual care (n=240) | 70.7 | 112 (70.9) | 1.04 (0.57, 1.89) |

| | **Pre-implementation cohort** M (SD) | **Post-implementation cohort** M (SD) | **Difference between pre-implementation [REF] and post-implementation cohorts** ALSM[1] (95% CI) |
|---|---|---|---|
| DECISION REGRET ABOUT INTENDED CONTRACEPTIVE METHOD(S) | | | |
| **Four weeks after the health care visit (n=1,250)** | | | |
| Video (n=260) | 0.82 (0.83) | 0.61 (0.62) | -0.16 (-0.35, 0.02) |
| Decision aids (n=232) | 0.59 (0.64) | 0.61 (0.62) | 0.00 (-0.19, 0.18) |
| Dual (n=337) | 0.56 (0.63) | 0.56 (0.67) | -0.03 (-0.19, 0.13) |
| Usual care (n=262) | 0.65 (0.60) | 0.56 (0.62) | -0.08 (-0.26, 0.10) |
| **Six months after the health care visit (n=1,250)** | | | |
| Video (n=237) | 0.66 (0.70) | 0.68 (0.77) | 0.04 (-0.15, 0.23) |
| Decision aids (n=215) | 0.70 (0.92) | 0.92 (0.90) | **0.21 (0.02, 0.40)** |
| Dual (n=275) | 0.66 (0.77) | 0.70 (0.78) | 0.01 (-0.17, 0.20) |
| Usual care (n=241) | 0.65 (0.77) | 0.65 (0.69) | 0.00 (-0.18, 0.19) |

*Notes.* ALSM = Adjusted Least Squares Mean, AOR = Adjusted Odds Ratio, CI = Confidence Interval, M = Mean, REF = Reference group, SD = Standard Deviation. [1]For the outcome of conversation about contraception, analyses adjusted for age, health insurance coverage, health literacy, educational attainment, ethnicity, race, number of pregnancies, number of births, number of abortions, number of miscarriages, and survey language. For the outcomes of satisfaction with conversation about contraception, intention to use one or more contraceptive method(s), intention to use highly effective contraceptive method, and values concordance immediately after the health care visit, analyses adjusted for age, health insurance coverage, health literacy, educational attainment, ethnicity, race, number of pregnancies, number of births, number of abortions, number of miscarriages, previous contraceptive method(s), and survey language. For the outcomes of values concordance four weeks and six months after the health care visit, use of intended contraceptive method(s), adherence to contraceptive method(s) used, satisfaction with contraceptive method(s), and decision regret, analyses adjusted for age, health insurance coverage, health literacy, educational attainment, ethnicity, race, number of pregnancies, number of births, number of abortions, number of miscarriages, and previous contraceptive method(s).

**Table A10.** Intervention effects on secondary outcomes: difference-in-differences analyses

| | **Difference-in-differences** AOR[1] (95% CI) | **P-value unadjusted for multiple comparisons** | **P-value adjusted for multiple comparisons** |
|---|---|---|---|
| CONVERSATION ABOUT CONTRACEPTION (n=4,865) | | | |
| Video vs. Usual care [REF] | 0.61 (0.37, 1.02) | 0.06 | 0.31 |
| Decision aids vs. Usual care [REF] | 1.12 (0.61, 2.03) | 0.72 | 0.99 |
| Dual vs. Video [REF] | 1.56 (0.94, 2.60) | 0.09 | 0.40 |
| Dual vs. Decision aids [REF] | 0.86 (0.47, 1.56) | 0.61 | 0.97 |
| ***Prespecified subgroup excluding those not at risk of unintended pregnancy (n=4,619)*** | | | |
| Video vs. Usual care [REF] | 0.60 (0.34, 1.04) | 0.07 | 0.34 |
| Decision aids vs. Usual care [REF] | 1.18 (0.60, 2.31) | 0.63 | 0.97 |
| Dual vs. Video [REF] | 1.22 (0.65, 2.29) | 0.53 | 0.94 |
| Dual vs. Decision aids [REF] | 0.62 (0.30, 1.29) | 0.20 | 0.65 |
| ***Prespecified subgroup excluding those not at risk of unintended pregnancy and those who reported that they did not want or need to talk about contraception (n=4,198)*** | | | |
| Video vs. Usual care [REF] | 0.66 (0.19, 2.25) | 0.50 | 0.93 |
| Decision aids vs. Usual care [REF] | 1.81 (0.30, 10.94) | 0.52 | 0.94 |
| Dual vs. Video [REF] | 1.60 (0.43, 5.92) | 0.48 | 0.92 |
| Dual vs. Decision aids [REF] | 0.58 (0.09, 3.70) | 0.57 | 0.95 |
| SATISFACTION WITH CONVERSATION ABOUT CONTRACEPTION (n=4,085) | | | |
| Video vs. Usual care [REF] | 1.10 (0.57, 2.12) | 0.78 | 0.99 |
| Decision aids vs. Usual care [REF] | 1.11 (0.53, 2.33) | 0.78 | 0.99 |
| Dual vs. Video [REF] | 0.85 (0.46, 1.59) | 0.62 | 0.97 |
| Dual vs. Decision aids [REF] | 0.84 (0.42, 1.70) | 0.63 | 0.97 |
| INTENDED CONTRACEPTIVE METHOD(S) (n=4,082) | | | |
| Video vs. Usual care [REF] | 0.98 (0.46, 2.09) | 0.96 | 1.00 |
| Decision aids vs. Usual care [REF] | 0.77 (0.36, 1.62) | 0.49 | 0.92 |
| Dual vs. Video [REF] | 0.61 (0.28, 1.31) | 0.20 | 0.65 |
| Dual vs. Decision aids [REF] | 0.78 (0.37, 1.66) | 0.52 | 0.94 |
| ***Prespecified subgroup excluding those not at risk of unintended pregnancy (n=3,865)*** | | | |
| Video vs. Usual care [REF] | 0.64 (0.23, 1.78) | 0.39 | 0.87 |
| Decision aids vs. Usual care [REF] | 0.47 (0.16, 1.38) | 0.17 | 0.60 |
| Dual vs. Video [REF] | 0.54 (0.18, 1.64) | 0.28 | 0.76 |
| Dual vs. Decision aids [REF] | 0.73 (0.24, 2.28) | 0.59 | 0.96 |
| INTENTION TO USE A HIGHLY EFFECTIVE CONTRACEPTIVE METHOD (n=4,082) | | | |
| Video vs. Usual care [REF] | 0.97 (0.56, 1.70) | 0.92 | 1.00 |
| Decision aids vs. Usual care [REF] | 1.05 (0.62, 1.77) | 0.85 | 1.00 |
| Dual vs. Video [REF] | 0.73 (0.42, 1.25) | 0.25 | 0.72 |
| Dual vs. Decision aids [REF] | 0.67 (0.41, 1.11) | 0.12 | 0.50 |
| ***Prespecified subgroup excluding those not at risk of unintended pregnancy (n=3,865)*** | | | |
| Video vs. Usual care [REF] | 0.88 (0.50, 1.57) | 0.67 | 0.98 |
| Decision aids vs. Usual care [REF] | 1.06 (0.62, 1.82) | 0.83 | 1.00 |
| Dual vs. Video [REF] | 0.76 (0.44, 1.32) | 0.32 | 0.81 |
| Dual vs. Decision aids [REF] | 0.63 (0.38, 1.06) | 0.08 | 0.39 |
| ***Prespecified subgroup excluding those not at risk of unintended pregnancy and those who reported that they did not want or need to use a contraceptive method (n=3,779)*** | | | |
| Video vs. Usual care [REF] | 0.95 (0.53, 1.70) | 0.85 | 1.00 |
| Decision aids vs. Usual care [REF] | 1.12 (0.64, 1.94) | 0.70 | 0.98 |
| Dual vs. Video [REF] | 0.72 (0.41, 1.26) | 0.25 | 0.73 |
| Dual vs. Decision aids [REF] | 0.61 (0.36, 1.03) | 0.07 | 0.34 |

| | | | |
|---|---|---|---|
| VALUES CONCORDANCE OF INTENDED CONTRACEPTIVE METHOD(S) | | | |
| **Immediately after the health care visit (n=3,688)** | | | |
| Video vs. Usual care [REF] | 1.04 (0.67, 1.59) | 0.87 | 1.00 |
| Decision aids vs. Usual care [REF] | 1.12 (0.73, 1.70) | 0.61 | 0.97 |
| Dual vs. Video [REF] | 1.15 (0.76, 1.75) | 0.50 | 0.93 |
| Dual vs. Decision aids [REF] | 1.07 (0.71, 1.61) | 0.74 | 0.99 |
| **Four weeks after the health care visit (n=1,252)** | | | |
| Video vs. Usual care [REF] | 0.72 (0.33, 1.56) | 0.40 | 0.87 |
| Decision aids vs. Usual care [REF] | 0.56 (0.26, 1.20) | 0.14 | 0.53 |
| Dual vs. Video [REF] | 0.97 (0.47, 2.02) | 0.93 | 1.00 |
| Dual vs. Decision aids [REF] | 1.25 (0.60, 2.58) | 0.55 | 0.95 |
| **Six months after the health care visit (n=1,252)** | | | |
| Video vs. Usual care [REF] | 0.66 (0.30, 1.47) | 0.31 | 0.80 |
| Decision aids vs. Usual care [REF] | 0.51 (0.23, 1.14) | 0.10 | 0.45 |
| Dual vs. Video [REF] | 1.42 (0.65, 3.12) | 0.38 | 0.86 |
| Dual vs. Decision aids [REF] | 1.84 (0.83, 4.07) | 0.13 | 0.51 |
| USE OF INTENDED CONTRACEPTIVE METHOD(S) | | | |
| **Four weeks after the health care visit (n=1,252)** | | | |
| Video vs. Usual care [REF] | 0.98 (0.45, 2.11) | 0.96 | 1.00 |
| Decision aids vs. Usual care [REF] | 0.83 (0.39, 1.79) | 0.64 | 0.97 |
| Dual vs. Video [REF] | 0.83 (0.40, 1.72) | 0.61 | 0.97 |
| Dual vs. Decision aids [REF] | 0.97 (0.47, 2.01) | 0.94 | 1.00 |
| **Six months after the health care visit (n=1,252)** | | | |
| Video vs. Usual care [REF] | 0.63 (0.29, 1.40) | 0.26 | 0.74 |
| Decision aids vs. Usual care [REF] | **0.45 (0.20, 0.99)** | **0.05** | 0.27 |
| Dual vs. Video [REF] | 1.46 (0.66, 3.21) | 0.35 | 0.83 |
| Dual vs. Decision aids [REF] | 2.07 (0.94, 4.55) | 0.07 | 0.35 |
| ADHERENCE TO CONTRACEPTIVE METHOD(S) USED | | | |
| **Four weeks after the health care visit (n=1,205)** | | | |
| Video vs. Usual care [REF] | 1.20 (0.44, 3.29) | 0.73 | 0.99 |
| Decision aids vs. Usual care [REF] | 0.43 (0.14, 1.31) | 0.14 | 0.53 |
| Dual vs. Video [REF] | 0.95 (0.37, 2.46) | 0.91 | 1.00 |
| Dual vs. Decision aids [REF] | 2.64 (0.92, 7.62) | 0.07 | 0.36 |
| **Six months after the health care visit (n=1,205)** | | | |
| Video vs. Usual care [REF] | 2.20 (0.75, 6.46) | 0.15 | 0.56 |
| Decision aids vs. Usual care [REF] | 0.42 (0.11, 1.63) | 0.21 | 0.67 |
| Dual vs. Video [REF] | 0.38 (0.13, 1.11) | 0.08 | 0.37 |
| Dual vs. Decision aids [REF] | 1.98 (0.52, 7.50) | 0.31 | 0.80 |
| SATISFACTION WITH CONTRACEPTIVE METHOD(S) USED | | | |
| **Four weeks after the health care visit (n=1,249)** | | | |
| Video vs. Usual care [REF] | 1.02 (0.46, 2.27) | 0.96 | 1.00 |
| Decision aids vs. Usual care [REF] | 0.70 (0.31, 1.55) | 0.38 | 0.86 |
| Dual vs. Video [REF] | 0.73 (0.34, 1.55) | 0.41 | 0.88 |
| Dual vs. Decision aids [REF] | 1.06 (0.50, 2.26) | 0.87 | 1.00 |
| **Six months after the health care visit (n=1,249)** | | | |
| Video vs. Usual care [REF] | 1.09 (0.46, 2.55) | 0.85 | 1.00 |
| Decision aids vs. Usual care [REF] | 0.79 (0.33, 1.85) | 0.58 | 0.96 |
| Dual vs. Video [REF] | **0.29 (0.11, 0.76)** | **0.01** | 0.10 |
| Dual vs. Decision aids [REF] | 0.40 (0.15, 1.06) | 0.06 | 0.33 |

| | Difference-in-differences ALSM[1] (95% CI) | P-value unadjusted for multiple comparisons | P-value adjusted for multiple comparisons |
|---|---|---|---|
| DECISION REGRET ABOUT INTENDED CONTRACEPTIVE METHOD(S) | | | |
| **Four weeks after the health care visit (n=1,250)** | | | |
| Video vs. Usual care [REF] | -0.08 (-0.34, 0.18) | 0.53 | 0.94 |
| Decision aids vs. Usual care [REF] | 0.08 (-0.18, 0.34) | 0.55 | 0.95 |
| Dual vs. Video [REF] | 0.14 (-0.11, 0.38) | 0.28 | 0.76 |
| Dual vs. Decision aids [REF] | -0.02 (-0.27, 0.22) | 0.84 | 1.00 |
| **Six months after the health care visit (n=1,250)** | | | |
| Video vs. Usual care [REF] | 0.04 (-0.23, 0.31) | 0.78 | 0.99 |
| Decision aids vs. Usual care [REF] | 0.21 (-0.06, 0.48) | 0.13 | 0.52 |
| Dual vs. Video [REF] | -0.03 (-0.29, 0.23) | 0.82 | 1.00 |
| Dual vs. Decision aids [REF] | -0.20 (-0.47, 0.07) | 0.14 | 0.54 |

*Notes.* ALSM = Adjusted Least Squares Mean, AOR = Adjusted Odds Ratio, CI = Confidence Interval, REF = Reference group. [1]For the outcome of conversation about contraception, analyses adjusted for age, health insurance coverage, health literacy, educational attainment, ethnicity, race, number of pregnancies, number of births, number of abortions, number of miscarriages, and survey language. For the outcomes of satisfaction with conversation about contraception, intention to use one or more contraceptive method(s), intention to use highly effective contraceptive method, and values concordance immediately after the health care visit, analyses adjusted for age, health insurance coverage, health literacy, educational attainment, ethnicity, race, number of pregnancies, number of births, number of abortions, number of miscarriages, previous contraceptive method(s), and survey language. For the outcomes of values concordance four weeks and six months after the health care visit, use of intended contraceptive method(s), adherence to contraceptive method(s) used, satisfaction with contraceptive method(s), and decision regret, analyses adjusted for age, health insurance coverage, health literacy, educational attainment, ethnicity, race, number of pregnancies, number of births, number of abortions, number of miscarriages, and previous contraceptive method(s).

#### APPENDIX 15 Supplementary results tables: process outcomes

**Table A11.** Patient-reported intervention exposure among pre-implementation and post-implementation cohorts

| | Video | Decision aids | Dual | Usual care |
|---|---|---|---|---|
| PRE-IMPLEMENTATION COHORT (n=1683) | (n=370) | (n=470) | (n=443) | (n=400) |
| **Video** | | | | |
| Watched whole video before visit | 6 (1.6) | 2 (0.4%) | 13 (3.0) | 4 (1.0) |
| Watch whole video during or after visit | 5 (1.4) | 7 (1.5%) | 6 (1.4) | 1 (0.3) |
| Watched some of video | 4 (1.1) | 1 (0.2%) | 7 (1.6) | 2 (0.5) |
| Watched none of video | 351 (95.9) | 458 (97.9%) | 413 (94.1) | 392 (98.2) |
| *Missing* | 4 | 2 | 4 | 1 |
| **Decision aids** | | | | |
| Looked at decision aid(s) during visit | 61 (16.7) | 86 (18.3) | 59 (13.5) | 39 (9.8) |
| Looked at decision aid(s) before or after visit | 72 (19.7) | 129 (27.5) | 101 (23.2) | 71 (17.9) |
| Did not look at decision aid(s) | 233 (63.7) | 254 (54.2) | 276 (63.3) | 287 (72.3) |
| *Missing* | 4 | 1 | 7 | 3 |
| **Video and decision aids** | | | | |
| Watched whole video before visit + looked at decision aid(s) during visit | 3 (0.8) | 1 (0.2) | 4 (0.9) | 1 (0.3) |
| Some other exposure to the video and decision aid(s) | 131 (35.8) | 216 (46.2) | 162 (37.2) | 111 (28.0) |
| Watched none of video + did not look at decision aid(s) | 232 (63.4) | 251 (53.6) | 270 (61.9) | 285 (71.8) |
| *Missing* | 4 | 2 | 7 | 3 |
| POST-IMPLEMENTATION COHORT (n=3335) | (n=875) | (n=671) | (n=945) | (n=844) |
| **Video** | | | | |
| Watched whole video while waiting | 160 (18.8) | 36 (5.6) | 162 (17.7) | 40 (5.0) |
| Watched some of video while waiting | 18 (2.1) | 14 (2.2) | 14 (1.5) | 18 (2.3) |
| Watched none of video while waiting | 674 (79.1) | 593 (92.2) | 737 (80.7) | 737 (92.7) |
| *Not sure or missing* | 23 | 28 | 32 | 49 |
| **Prompt card** | | | | |
| Received prompt card while waiting | 156 (19.3) | 140 (23.9) | 238 (27.1) | 74 (9.5) |
| Did not receive prompt card while waiting | 652 (80.7) | 446 (76.1) | 640 (72.9) | 702 (90.5) |
| *Not sure or missing* | 67 | 85 | 67 | 68 |
| **Video and prompt card** | | | | |
| Watched whole video while waiting + received prompt card while waiting | 75 (9.4) | 23 (4.0) | 74 (8.6) | 16 (2.1) |
| Watched whole video while waiting + did not receive prompt card while waiting | 74 (9.3) | 7 (1.2) | 75 (8.7) | 16 (2.1) |
| Watched some of video while waiting + received prompt card while waiting | 7 (0.9) | 7 (1.2) | 4 (0.5) | 4 (0.5) |
| Watched some of video while waiting + did not receive prompt card while waiting | 7 (0.9) | 5 (0.9) | 8 (0.9) | 14 (1.9) |
| Watched none of video while waiting + received prompt card while waiting | 69 (8.7) | 101 (17.6) | 149 (17.3) | 48 (6.4) |
| Watched none of video while waiting + did not receive prompt card while waiting | 564 (70.9) | 431 (75.1) | 552 (64.0) | 651 (86.9) |
| *Not sure or missing* | 79 | 97 | 83 | 95 |
| **Decision aids** | | | | |
| Used decision aid(s) with provider | 71 (9.0) | 194 (31.5) | 147 (16.9) | 94 (12.0) |
| Did not use decision aid(s) with provider but were given | 146 (18.5) | 158 (25.6) | 194 (22.3) | 143 (18.2) |
| Did not use decision aid(s) with provider and were not given | 571 (72.5) | 264 (42.9) | 528 (60.8) | 548 (69.8) |
| *Not sure or missing* | 87 | 55 | 76 | 59 |
| **Video, prompt card, and decision aids** | | | | |
| Watched whole video while waiting + received prompt card while waiting + used decision aid(s) with provider | 26 (3.5) | 17 (3.2) | 41 (5.0) | 9 (1.3) |
| Some other exposure to the video, prompt card, and decision aid(s) | 268 (36.3) | 294 (54.9) | 382 (47.0) | 223 (31.3) |
| Watched none of video while waiting + did not receive prompt card while waiting + did not use decision aid(s) with provider and were not given | 444 (60.2) | 225 (42.0) | 389 (47.9) | 481 (67.5) |

| | | | | |
|---|---|---|---|---|
| *Not sure or missing* | 137 | 135 | 133 | 131 |

*Notes*. Grey cells signify intended exposure. Information on variable coding is available on request.

**Table A12.** Acceptability of interventions among patients exposed as intended in the post-implementation cohort

| | **Would recommend to a friend** | | **Would not recommend to a friend** | |
|---|---|---|---|---|
| | Freq. | % (95% CI) | Freq. | % (95% CI) |
| Watched whole video while waiting (n=322) | 305 | 94.7 (91.7, 96.9) | 17 | 5.3 (3.1, 8.3) |
| Received prompt card while waiting (n=394) | 360 | 91.4 (88.2, 94.0) | 34 | 8.6 (6.0, 11.8) |
| Used decision aid(s) with provider (n=341) | 336 | 98.5 (96.6, 99.5) | 5 | 1.5 (0.5, 3.4) |
