## Supplementary material for "Right For Me: a pragmatic multi-arm cluster randomised controlled trial of two interventions for increasing shared decision-making about contraceptive methods": Reporting Checklist

### Table 1: CONSORT 2010 checklist of information to include when reporting a cluster randomised trial

| Section/Topic | Item No | Standard Checklist item | Extension for cluster designs | Page No * |
|---|---|---|---|---|
| **Title and abstract** | | | | |
| | 1a | Identification as a randomised trial in the title | Identification as a cluster randomised trial in the title | p1 |
| | 1b | Structured summary of trial design, methods, results, and conclusions (for specific guidance see CONSORT for abstracts)[1,2] | See table 2 | p2 |
| **Introduction** | | | | |
| **Background and objectives** | 2a | Scientific background and explanation of rationale | Rationale for using a cluster design | p3 + Study protocol p2 |
| | 2b | Specific objectives or hypotheses | Whether objectives pertain to the the cluster level, the individual participant level or both | p3 + Appendix pp2-3 |
| **Methods** | | | | |
| **Trial design** | 3a | Description of trial design (such as parallel, factorial) including allocation ratio | Definition of cluster and description of how the design features apply to the clusters | pp4-5 |
| | 3b | Important changes to methods after trial commencement (such as eligibility criteria), with reasons | | pp9-12 + Appendix p101 |
| **Participants** | 4a | Eligibility criteria for participants | Eligibility criteria for clusters | pp4-5 |
| | 4b | Settings and locations where the data were collected | | pp4-5 |
| **Interventions** | 5 | The interventions for each group with sufficient details to allow replication, | Whether interventions pertain to the cluster level, the individual participant | pp6-7 + p22 + Appendix pp11-94 + Appendix |

| | | | | |
|---|---|---|---|---|
| | | including how and when they were actually administered | level or both | pp111-112 |
| **Outcomes** | 6a | Completely defined pre-specified primary and secondary outcome measures, including how and when they were assessed | Whether outcome measures pertain to the cluster level, the individual participant level or both | pp7-10 |
| | 6b | Any changes to trial outcomes after the trial commenced, with reasons | | pp9-12 + Appendix p101 |
| **Sample size** | 7a | How sample size was determined | Method of calculation, number of clusters(s) (and whether equal or unequal cluster sizes are assumed), cluster size, a coefficient of intracluster correlation (ICC or *k*), and an indication of its uncertainty | p10 + Study protocol pp8-9 |
| | 7b | When applicable, explanation of any interim analyses and stopping guidelines | | pp4-5 |
| **Randomisation:** | | | | |
| **Sequence generation** | 8a | Method used to generate the random allocation sequence | | p5 |
| | 8b | Type of randomisation; details of any restriction (such as blocking and block size) | Details of stratification or matching if used | p5 |
| **Allocation concealment mechanism** | 9 | Mechanism used to implement the random allocation sequence (such as sequentially numbered containers), describing any steps taken to conceal the sequence until interventions were assigned | Specification that allocation was based on clusters rather than individuals and whether allocation concealment (if any) was at the cluster level, the individual participant level or both | p5 |

| | | | | |
|---|---|---|---|---|
| **Implementation** | 10 | Who generated the random allocation sequence, who enrolled participants, and who assigned participants to interventions | Replace by 10a, 10b and 10c | p5 |
| | 10a | | Who generated the random allocation sequence, who enrolled clusters, and who assigned clusters to interventions | p5 |
| | 10b | | Mechanism by which individual participants were included in clusters for the purposes of the trial (such as complete enumeration, random sampling) | pp5-6 |
| | 10c | | From whom consent was sought (representatives of the cluster, or individual cluster members, or both), and whether consent was sought before or after randomisation | pp5-6 + Study protocol p10 |
| **Blinding** | 11a | If done, who was blinded after assignment to interventions (for example, participants, care providers, those assessing outcomes) and how | | p5 |
| | 11b | If relevant, description of the similarity of interventions | | pp6-7 |
| **Statistical methods** | 12a | Statistical methods used to compare groups for primary and secondary outcomes | How clustering was taken into account | pp10-12 |
| | 12b | Methods for additional analyses, such as subgroup analyses and adjusted analyses | | pp10-12 |

| Results | | | | |
|---|---|---|---|---|
| **Participant flow (a diagram is strongly recommended)** | 13a | For each group, the numbers of participants who were randomly assigned, received intended treatment, and were analysed for the primary outcome | For each group, the numbers of clusters that were randomly assigned, received intended treatment, and were analysed for the primary outcome | pp12-14 + Appendix p102 |
| | 13b | For each group, losses and exclusions after randomisation, together with reasons | For each group, losses and exclusions for both clusters and individual cluster members | pp12-14 + Appendix pp96-99 + Appendix p102 |
| **Recruitment** | 14a | Dates defining the periods of recruitment and follow-up | | p4 |
| | 14b | Why the trial ended or was stopped | | p4 |
| **Baseline data** | 15 | A table showing baseline demographic and clinical characteristics for each group | Baseline characteristics for the individual and cluster levels as applicable for each group | p16 |
| **Numbers analysed** | 16 | For each group, number of participants (denominator) included in each analysis and whether the analysis was by original assigned groups | For each group, number of clusters included in each analysis | p10 + p14 + Appendix pp96-99 + Appendix p102 |
| **Outcomes and estimation** | 17a | For each primary and secondary outcome, results for each group, and the estimated effect size and its precision (such as 95% confidence interval) | Results at the individual or cluster level as applicable and a coefficient of intracluster correlation (ICC or k) for each primary outcome | pp19-22 + Appendix pp103-110 |
| | 17b | For binary outcomes, presentation of both absolute and relative effect sizes is recommended | | pp19-22 + Appendix pp103-110 |
| **Ancillary analyses** | 18 | Results of any other analyses performed, | | pp19-22 + Appendix |

| | | | | |
|---|---|---|---|---|
| | | including subgroup analyses and adjusted analyses, distinguishing pre-specified from exploratory | | pp103-110 |
| **Harms** | 19 | All important harms or unintended effects in each group (for specific guidance see CONSORT for harms[3]) | | pp19-22 + Appendix pp103-110 |
| **Discussion** | | | | |
| **Limitations** | 20 | Trial limitations, addressing sources of potential bias, imprecision, and, if relevant, multiplicity of analyses | | p23 |
| **Generalisability** | 21 | Generalisability (external validity, applicability) of the trial findings | Generalisability to clusters and/or individual participants (as relevant) | p23 |
| **Interpretation** | 22 | Interpretation consistent with results, balancing benefits and harms, and considering other relevant evidence | | pp22-24 |
| **Other information** | | | | |
| **Registration** | 23 | Registration number and name of trial registry | | p2 |
| **Protocol** | 24 | Where the full trial protocol can be accessed, if available | | p3 |
| **Funding** | 25 | Sources of funding and other support (such as supply of drugs), role of funders | | p28 |

** Note: page numbers optional depending on journal requirements*

#### Table 2: Extension of CONSORT for abstracts[1,2] to reports of cluster randomised trials

| Item | Standard Checklist item | Extension for cluster trials |
|---|---|---|
| **Title** | Identification of study as randomised | **Identification of study as cluster randomised** |
| **Trial design** | Description of the trial design (e.g. parallel, cluster, non-inferiority) | |
| **Methods** | | |
| **Participants** | Eligibility criteria for participants and the settings where the data were collected | **Eligibility criteria for clusters** |
| **Interventions** | Interventions intended for each group | |
| **Objective** | Specific objective or hypothesis | **Whether objective or hypothesis pertains to the cluster level, the individual participant level or both** |
| **Outcome** | Clearly defined primary outcome for this report | **Whether the primary outcome pertains to the cluster level, the individual participant level or both** |
| **Randomization** | How participants were allocated to interventions | **How clusters were allocated to interventions** |
| **Blinding (masking)** | Whether or not participants, care givers, and those assessing the outcomes were blinded to group assignment | |
| **Results** | | |
| **Numbers randomized** | Number of participants randomized to each group | **Number of clusters randomized to each group** |
| **Recruitment** | Trial status[1] | |
| **Numbers analysed** | Number of participants analysed in each group | **Number of clusters analysed in each group** |
| **Outcome** | For the primary outcome, a result for each group and the estimated effect size and its precision | **Results at the cluster or individual participant level as applicable for each primary outcome** |
| **Harms** | Important adverse events or side effects | |
| **Conclusions** | General interpretation of the results | |
| **Trial registration** | Registration number and name of trial register | |
| **Funding** | Source of funding | |

[1] Relevant to Conference Abstracts
